# Self-Supervised Learning for Biomedical Signal Processing: A Systematic Review on ECG and PPG Signals

**DOI:** 10.1101/2024.09.30.24314588

**Authors:** Cheng Ding, Chenwei Wu

## Abstract

Self-supervised learning has emerged as a promising paradigm for enhancing the analysis of physiological signals, particularly Electrocardiogram (ECG) and Photoplethysmogram (PPG) data. This review paper surveys the application of self-supervised learning techniques in the domain of ECG and PPG signal analysis. Traditional supervised methods often rely on labeled data, which can be limited and costly to acquire in medical contexts. Self-supervised learning leverages the inherent structure and temporal dependencies within ECG and PPG signals to train models without explicit annotations. By exploiting pretext tasks such as predicting time intervals, missing samples, or temporal order, self-supervised approaches can learn meaningful representations that capture crucial information for subsequent downstream tasks. This paper provides an overview of key self-supervised methods applied to ECG and PPG data, highlighting their advantages and challenges. Furthermore, it discusses the transferability of learned representations to various clinical applications, including arrhythmia detection, anomaly detection, and heart rate variability analysis. Through this comprehensive review, we shed light on the potential of self-supervised learning to revolutionize ECG and PPG signal processing, ultimately contributing to improved healthcare diagnostics and monitoring.

## 1 Introduction

Biomedical signals, acquired from the intricate depths of the human body cells, organs, and molecules, serve as profound indicators of our physiological existence. They embody a symphony of life, revealing themselves through various forms of signal processing. For instance, the ethereal EEG unravels the mind’s electrical whispers, while the captivating ECG unveils the rhythmic dance of the heart. The enchanting PPG reveals the pulsating rhythm of blood flow.

It unveils hidden patterns and intricacies. Through meticulous analysis, uncovers the pulse waveform, assesses arterial stiffness, and explores the realm of blood oxygenation. These captivating signals, initially harnessed for diagnosis and detection, have found their way into the realm of medical care, offering insights into the intricate workings of biological systems. Guided by the artistry of Artificial Intelligence, they undergo a metamorphosis, shedding noise, extracting exquisite features, unraveling the true essence of signal models, reducing dimensions to reveal dysfunctions and even unveiling glimpses of future pathologies and functional events.

The manual interpretation of ECG, though essential, is a time-consuming endeavor. Moreover, the intricate nature of complicated cases often leads to differing opinions among cardiologists. Given the vast scale, quality, and complexity of these data, relying solely on human experts or expert systems proves to be of limited utility, urging the need for data-driven methods to emerge. Machine learning techniques have been successfully utilized to detect cardiac abnormalities and identify sleep disorders. A groundbreaking development in this field comes in the form of deep learning (DL) models, particularly deep neural networks (DNNs), which have exhibited comparable or even superior performance to clinical cardiologists across various biomedical signal analysis tasks. When compared with classical machine learning approaches, supervised deep learning methods showcase outstanding advantages when it comes to modeling ECG and PPG data.

Supervised biomedical signal processing, while widely employed, poses various challenges that hinder its effectiveness and applicability in the field of healthcare. One of the key hurdles lies in the acquisition and annotation of labeled data, which can be time-consuming, resource-intensive, and subject to inter-observer variability. Additionally, the scarcity of annotated datasets in certain medical domains limits the ability to train accurate and robust models. These challenges have prompted researchers to explore alternative approaches such as self-supervised learning in order to leverage the vast amounts of unlabeled data available. Several studies have highlighted these challenges and emphasized the need for innovative solutions. For instance, the work of Hannun et al. [1] and Ribeiro et al. [2] delves into the time-consuming nature of manual ECG interpretation and the potential disagreements among cardiologists in complex cases. Similarly, Zhang et al. [3], Minchole et al. [4], and Al-Zaiti et al. [5] emphasize the limitations of supervised methods in detecting cardiac abnormalities, while Zarei and Asl. [6] and Pan et al. [7] discuss the challenges of labeled data scarcity in sleep disorder analysis. These papers shed light on the challenges faced in supervised biomedical signal processing and provide a foundation for exploring alternative approaches like self-supervised learning.

As a resolution a remarkable approach known as self-supervised learning has emerged, offering promising possibilities for unlocking hidden knowledge and patterns within physiological data. This innovative paradigm allows machines to learn directly from the inherent structure and relationships present in the signals themselves, without relying on labeled data. Self-supervised learning has gained significant attention in the field, particularly in the analysis of vital signals such as ECG and PPG. By harnessing the power of unsupervised learning, self-supervised methods aim to unveil intricate insights, enabling accurate diagnosis, disease detection, and personalized healthcare. The purpose of this survey paper is to provide an all-encompassing review and concise summary of the existing literature pertaining to self-supervised biomedical signal processing techniques, specifically focusing on ECG and PPG signals, within the realm of artificial intelligence. The paper aims to highlight the immense potential of self-supervised algorithms that leverage ECG and PPG signals in predicting and diagnosing cardiac diseases. Its objectives encompass a thorough examination of the methodologies employed in the literature, an evaluation of their performance metrics, an identification of ongoing challenges, and the proposition of potential solutions for future advancements in this field.

The intended audience for this extensive review spans across various domains, including researchers and practitioners entrenched in the realms of artificial intelligence, machine learning, and healthcare. Furthermore, it targets medical professionals, esteemed cardiologists, experts in biomedical informatics, and computational scientists engrossed in the intricate analysis of biomedical signals. The value of this survey extends to academia, students, decision-makers within the healthcare sphere, industry professionals specializing in healthcare technology, and researchers in related fields such as precision medicine and cardiology. Moreover, it captivates the attention of the broader scientific community fascinated by the convergence of AI, digital pathology, and the prognosis of heart disease risk. To facilitate comprehension, a comprehensive table (Table 1) accompanies the survey, elucidating commonly used terminologies and their respective descriptions.

**Table 1:** List of accronyms and abbreviations used in paper.

| Acronyms | Words | Acronyms | Words |
| --- | --- | --- | --- |
| ECG | Electrocardiogram | PPG | Photoplethysmography |
| SSL | Self Supervised Learning | DBN | Deep Belief Network |
| CNN | Convolutional Neural Network | HRV | Heart rate variability |
| PRV | Pulse rate variability | SpO2 | Oxygen saturation |
| QRS | Depolarization of ventricles | VAEs | Variational autoencoders |
| RNN | Recurrent Neural Network | LSTM | Long short term memory |
| GAN | Generative Adversarial network | EEG | Electroencephalogram |
| EMG | Electromyography | AI | Artificial Intelligence |
| DNN | Deep Neural Network |  |  |

### 1.1 Scope of the Review

In this captivating review article, we aim to offer a comprehensive and enchanting overview of the current research landscape surrounding the use of self-supervised learning in biomedical signal processing, with a primary focus on ECG and PPG signals. Our objective is to summarize the methodologies employed, address the challenges faced, and explore the advancements made in this field. By doing so, we seek to guide and inspire future research and development efforts that can enhance prognosis and treatment outcomes for patients.

To ensure the inclusiveness of our review, we conducted an extensive literature search across reputable platforms such as Elsevier, Springer, IEEE Xplore, and Google Scholar. Our search encompassed peer-reviewed journals and conferences, and we examined publications from the period between 2019 and 2023. Each selected paper’s adopted methodology underwent a comprehensive review process. Throughout our exploration, we not only emphasized the captivating approaches described in the literature but also shed light on areas that warrant further research and potential avenues for future investigations.

Remarkably, our investigation revealed a notable gap in existing reviews specifically focusing on self-supervised learning techniques in biomedical signal processing, including ECG and PPG. Therefore, this review holds the potential of being the pioneering work in this domain, providing an exceptional opportunity to bridge the gap between the computational community and medical specialists. Our hope is that this bridging will foster collaborative research and development endeavors in the application of self-supervised learning techniques for ECG and PPG analysis.

The interdisciplinary nature of this review makes it highly relevant and intriguing for a diverse range of professionals. Our target audience includes AI practitioners, medical statisticians, and forward-thinking cardiologists who possess a keen interest in implementing AI-driven solutions within clinical practice. Furthermore, we have taken care to describe publicly available ECG and PPG datasets that can facilitate biomedical signal analysis.

Ultimately, the grand aspiration of this review article is to provide valuable insights and guidance that can shape future research directions in the captivating field of biomedical signal processing. By doing so, we aspire to contribute to the ongoing progress and innovation in this domain, thereby benefiting both the scientific community and patients alike.

## 2 Survey Methodology

In this section, we have examined the criteria utilized for selecting literature for extracting relevant research articles in this review paper. Furthermore, we have exemplified the categorization of the reviewed articles according to their publication avenues including conferences and journals and the respective years in which they were published.

### 2.1 Papers Selection

In this section, we have examined the criteria utilized for selecting literature for extracting relevant research articles in this review paper. Furthermore, we have exemplified the categorization of the reviewed articles according to their publication avenues including conferences and journals and the respective years in which they were published. In this review paper, we have utilized PRISMA (Preferred Reporting Items for Systematic Reviews and Meta-Analyses) to present in detail the main data of the research articles included. PRISMA diagram is attached in Figure 1 below, it provides a conclusive view of the paper selection process. Our search for relevant papers was conducted on Elsevier, Springer, IEEE Xplore, and Google Scholar. Initially, a total of 7,860 papers were identified through search queries. Among these 7860 papers, unfortunately, we excluded a total of 5000 papers because they are totally irrelevant to the topic of research which is biomedical signal processing also the data of those papers are not based on either EEG or PPG, not focussed on selfsupervised learning or duplication. After screening the remaining 2,860 papers, we carefully selected 305 articles that met our criteria. The remaining 2,555 papers were found ineligible for this review as they did not fulfill the relevancy criteria. Specifically, some of these papers solely focused on other biomedical signals, rather than exploring ECG and PPG signal processing techniques based on self-supervised learning.

**Fig. 1:**
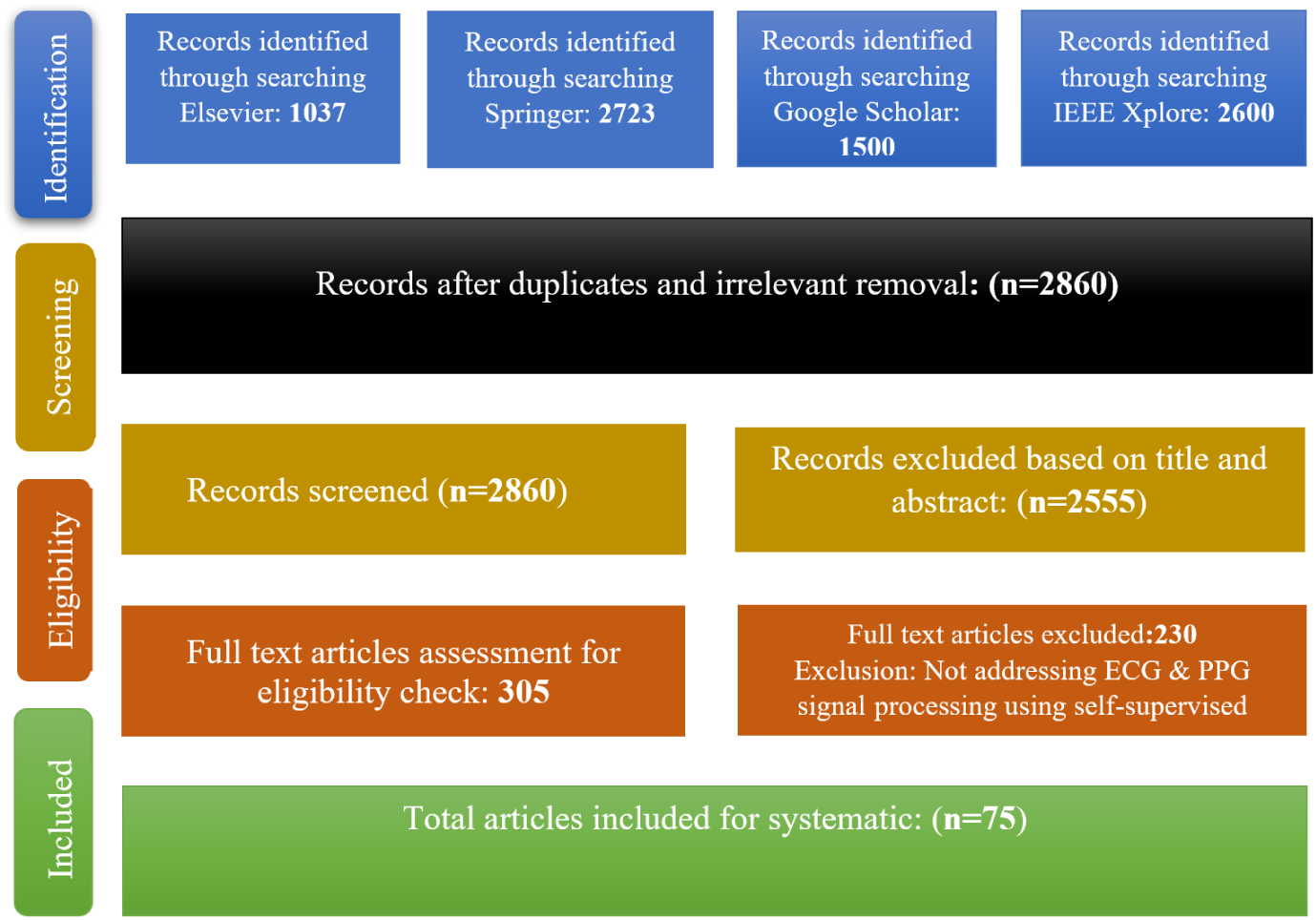
PRISMA - flow diagram of selected research articles for the review

Furthermore, out of the 305 selected articles, we conducted an additional screening for legitimacy and excluded 230 papers. These papers either focused on extracting features other than histological features (such as gene mutation and protein biomarkers) or employed techniques other than deep learning or machine learning, such as relying on expert pathologists for problem resolution. In the end, the total number of articles remaining for the systematization is equivalent to the number of 75.

### 2.2 Papers Extraction Methods

Relevant various platforms are used in the search of articles for this review paper, platforms used include Elsevier, Springer, IEEE Xplore, and Google Scholar. Mostly conference papers and peer-reviewed document papers are selected. Different keywords related to this review are used to search exactly relevant results. Keywords are diverse and it is combined with two logical operators which are “OR” and “AND”, both are used to get good results especially relevant to this review.

The list of keywords used in searching are:

- Biomedical signals, ECG, Self-supervised learning.
- PPG, Representation learning; Neural networks, Self-supervised learning
- Photoplethysmography, Electrocardiography, Transformer based
- ECG-based, Mental stress detection, discriminative Clustering, Adversarial Domain Adaptation.
- Electrocardiography, Transformers, Physiology, Pattern recognition
- Skin, Synchronization, Optical sensors, Heart rate; Wrist, Annotations, Photoplethysmography.

Using those keywords, the word cloud is generated. The extracted keywords taken from the research articles relating to this review are shown in Fig 2 The word cloud indicates the most used words in the research articles chosen for the review paper self-supervised learning for biomedical signal processing. The main goal is to transform into a sequence of all the available literature documents related to biomedical signal processing using ecg and ppg signals. So, if we look at the word cloud it highlights the main terms used which include neutral, physiology, deep signal, electrocardiography, synchronization, extractions, annotations, and many more.

**Fig. 2:**
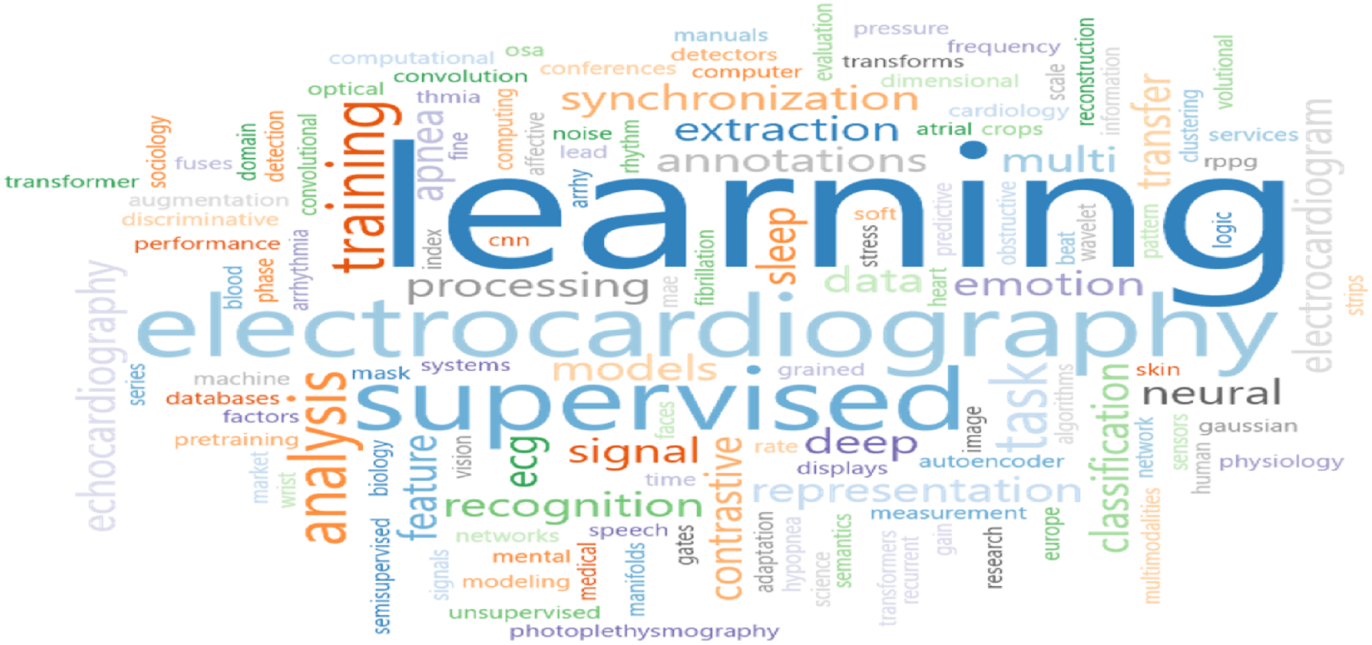
A picture of the most repeated keywords from research articles under review.

Paper inclusion and exclusion criteria are in detail described in Table 2. In the initial stage selection of papers is done based on their titles. If the titles do not fulfill the exclusion and inclusion criteria, in the later stage, the final selection is done based on what is in the abstract, the conclusion of the paper, and the paper’s model diagram.

**Table 2:**
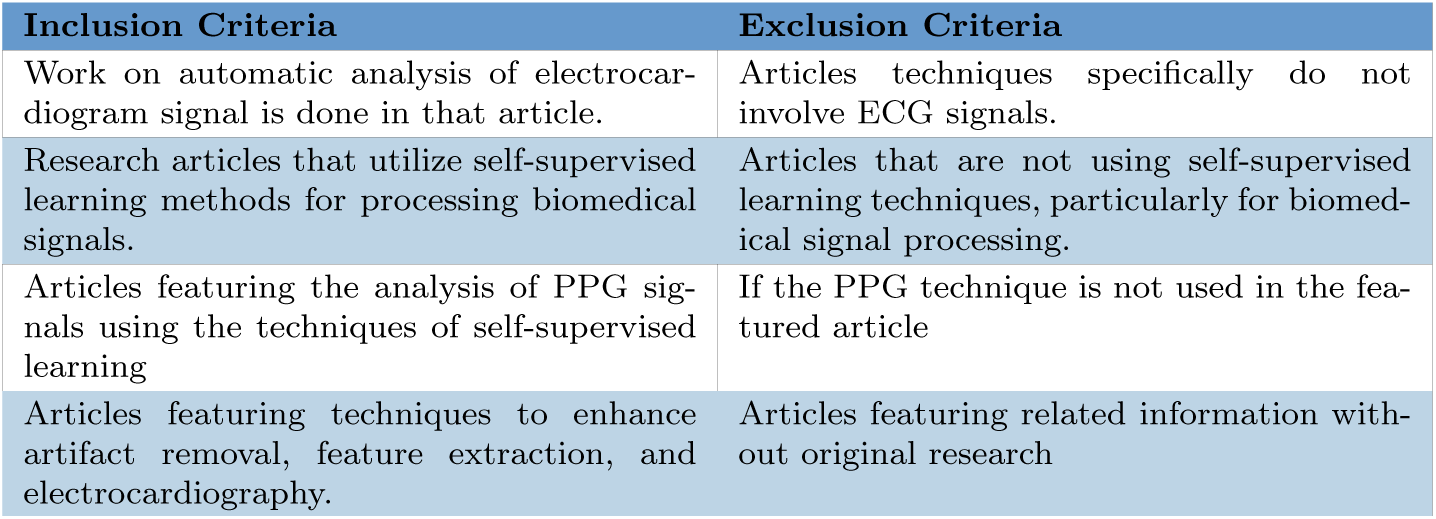
Inclusion Exclusion Criteria.

### 2.3 Papers dispersal

In this section, we present the distribution of the published papers among various journals, conferences, and years. The objective of this section is to offer a summary of the articles included in this review, demonstrating the extent of available literature in peer-reviewed publications and conferences, as well as the influence of the work.

Figure 3 shows the distribution of reviewed articles across the years.

**Fig. 3:**
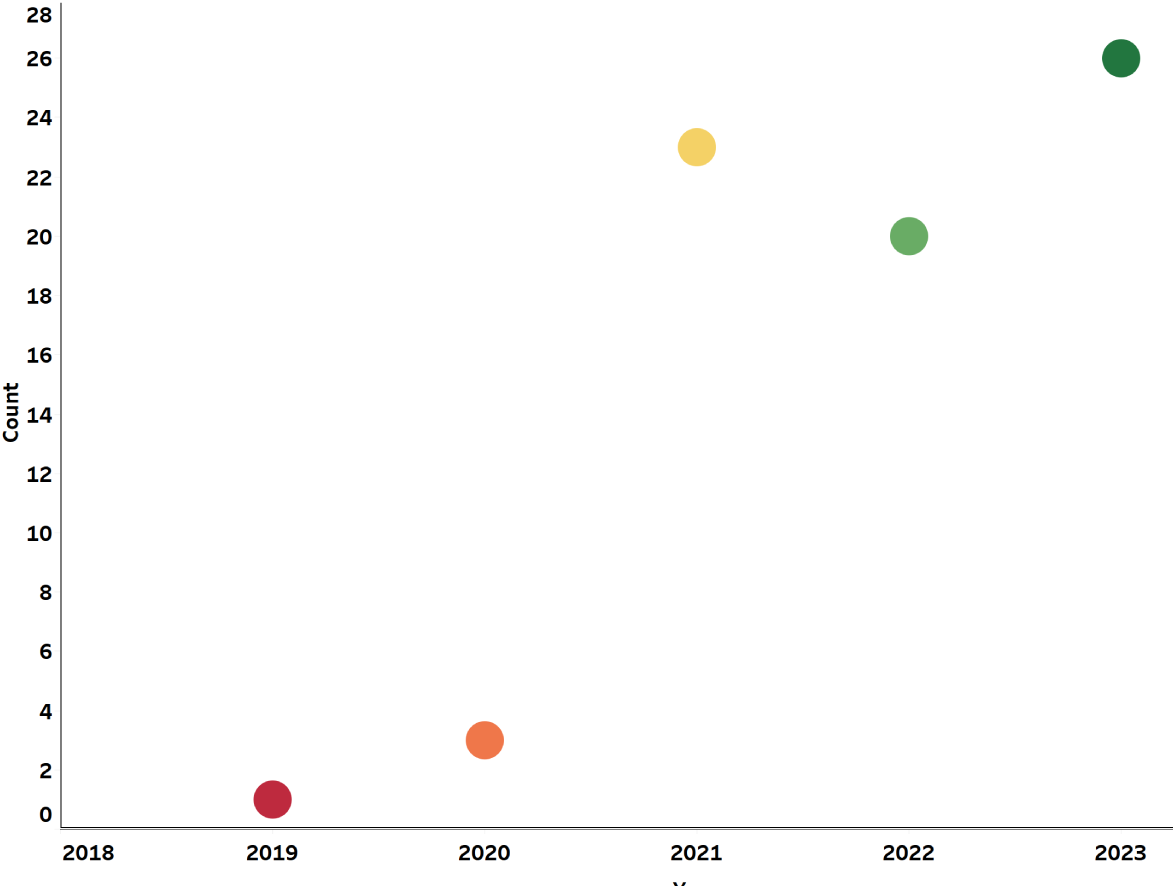
Bubble graph representing number of articles chosen from top ranked journals and conferences

The graph illustrates the progressive expansion of literature on ECG and PPG from 2019 to 2023. The research on biomedical signal processing using self supervised learning using ECG and PPG signals has shown consistent growth starting in 2019, with a significant surge in publications during 2023, with 26 research articles being published.

Figure 4 illustrates the distribution of papers according to their sources. It displays the count of self-supervised learning articles based on ECG and PPG data, extracted from various reputable journals and conferences. Each bubble in the figure represents the number of papers obtained from a specific journal or conference.

**Fig. 4:**
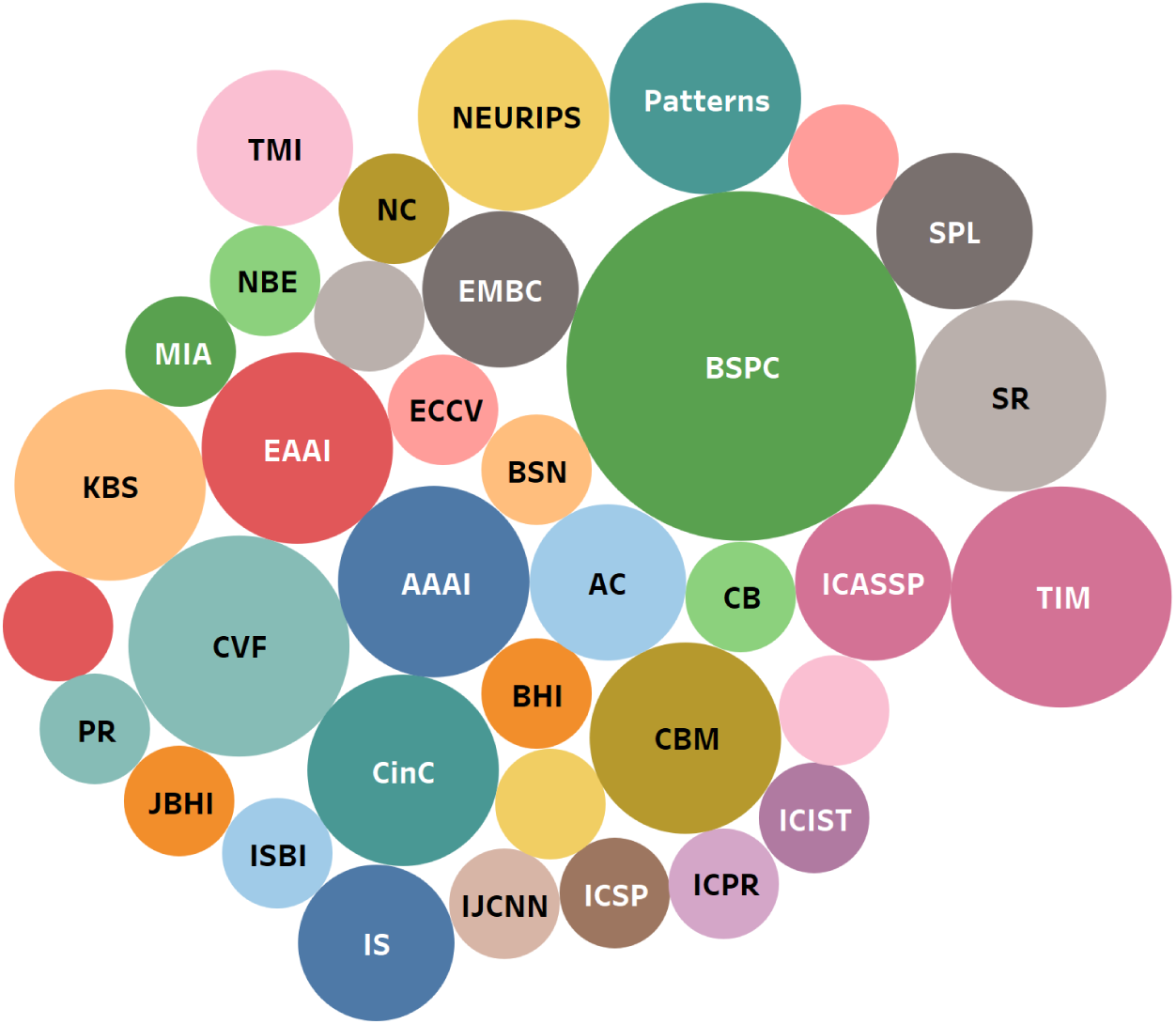
Bubble graph representing number of articles chosen from top ranked journals and conferences

**Fig. 5:**
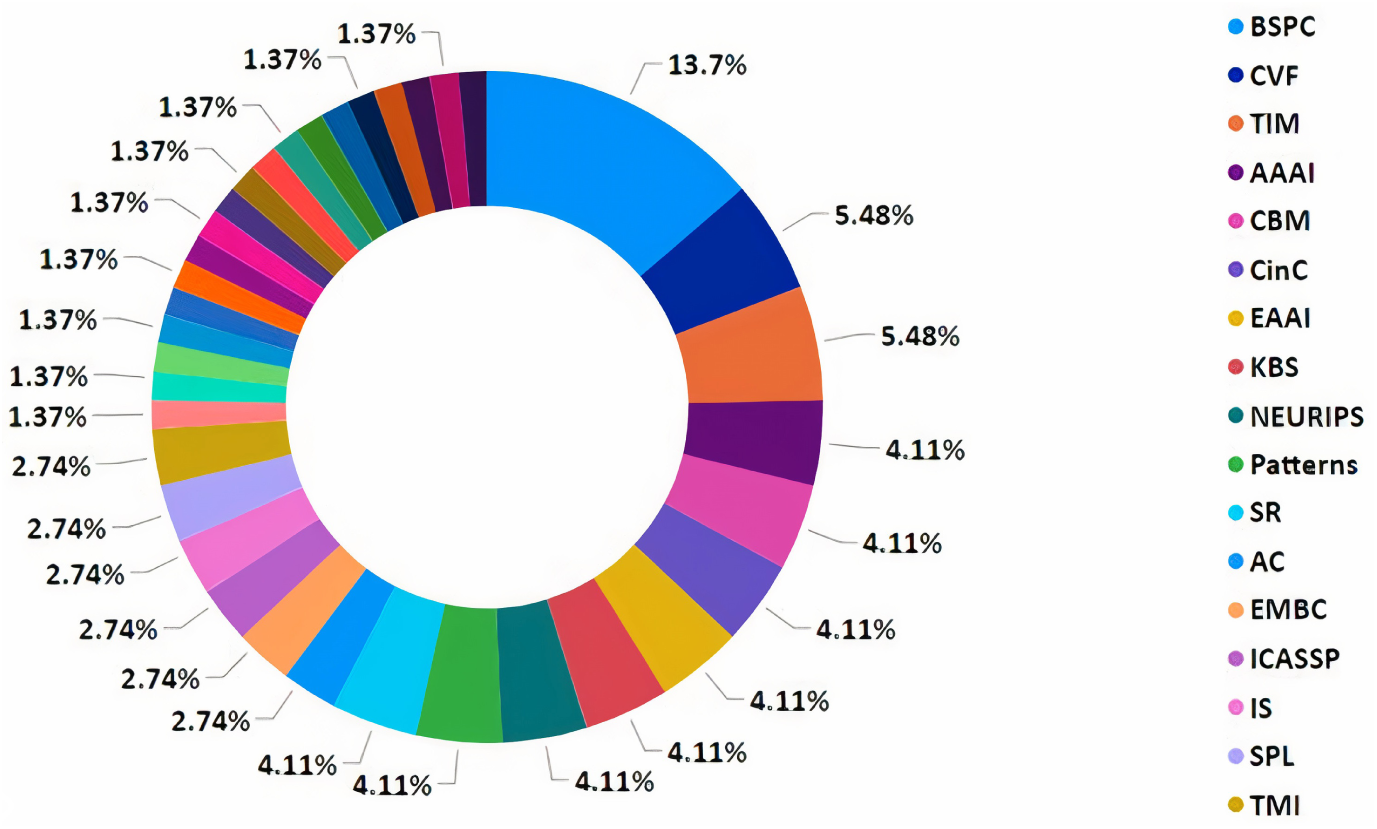
Bubble graph representing number of articles chosen from top ranked journals and conferences

**Fig. 6:**
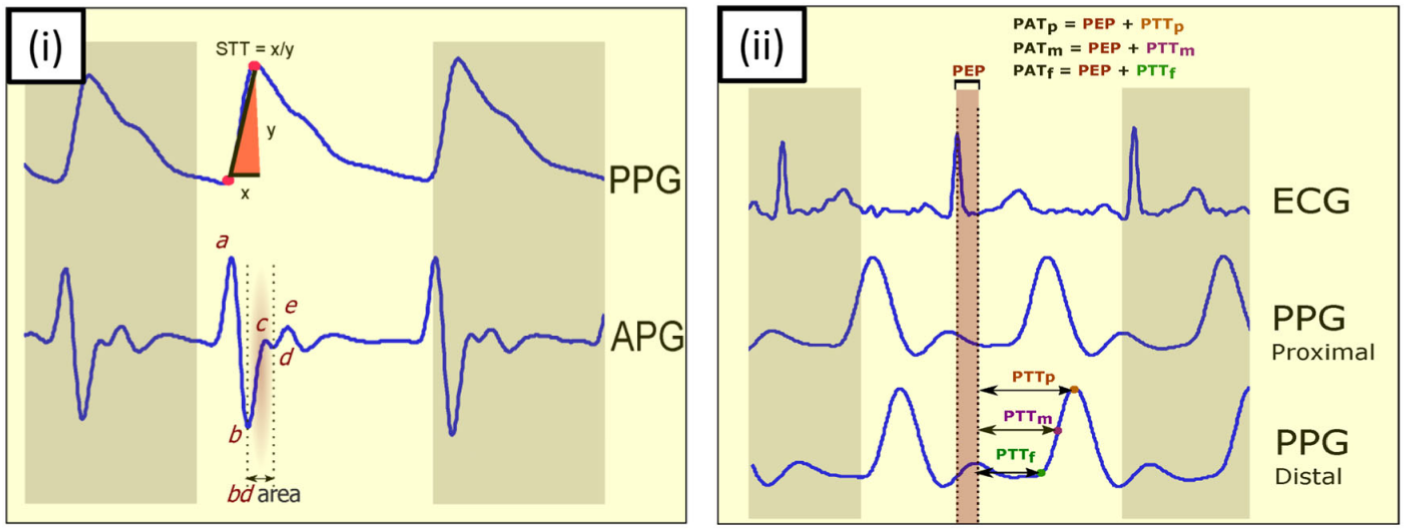
Sample ECG Signal

**Fig. 7:**
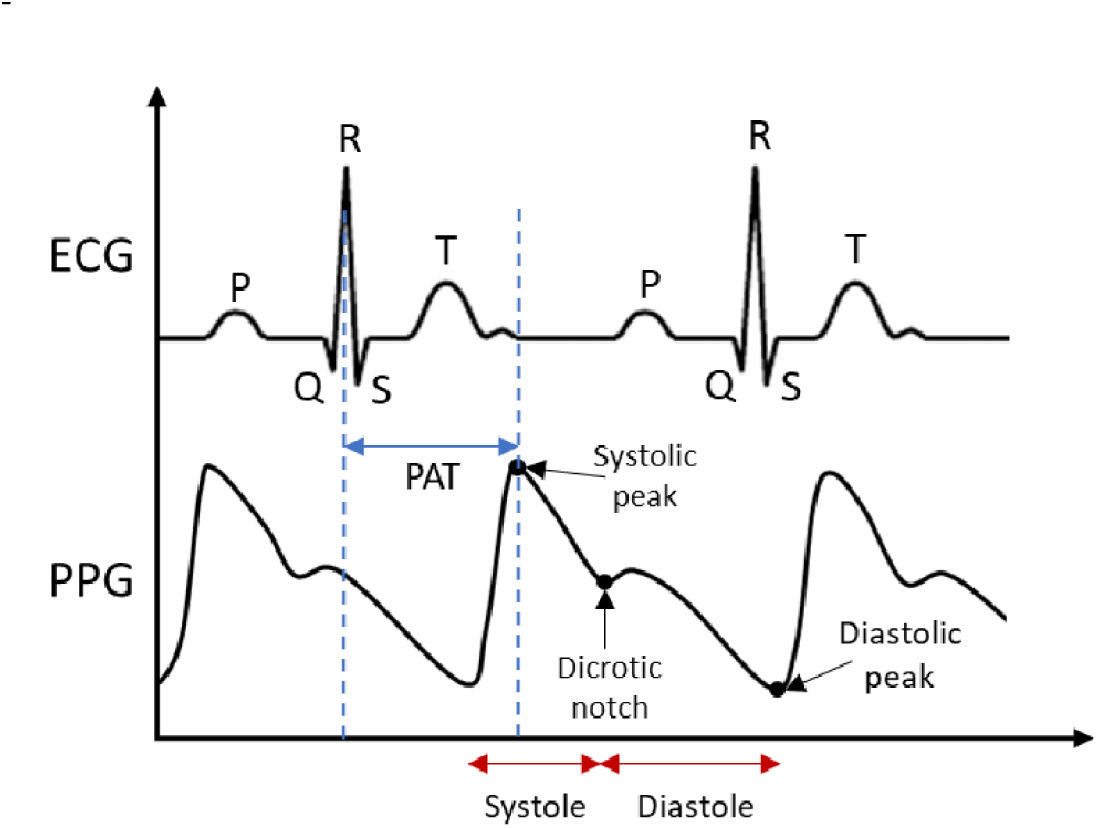
Illustrations depicting the schematic representations of ECG and PPG waveforms

**Fig. 8:**
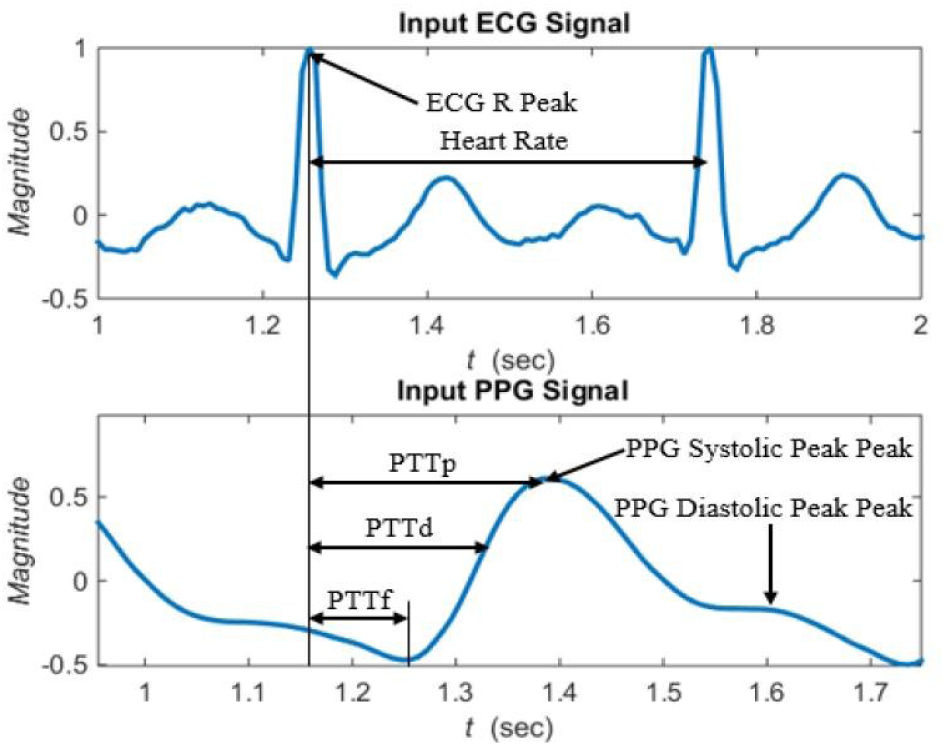
Sample ECG and PPG signal illustrating PPT calculation from R peaks of ECG signal and some point of PPG signal

The number of research articles focusing on self supervised learning for ECG and PPG in different journals and conferences is visually presented in Figure 9. Each venue percentage is depicted in the pie chart that corresponds to a specific segment color of the total block. The graphical representation indicates that the largest proportion of papers, totaling 13% articles, originated from BSPC journal. Moreover, CVF contributed 5% papers to the overall review.

**Fig. 9:**
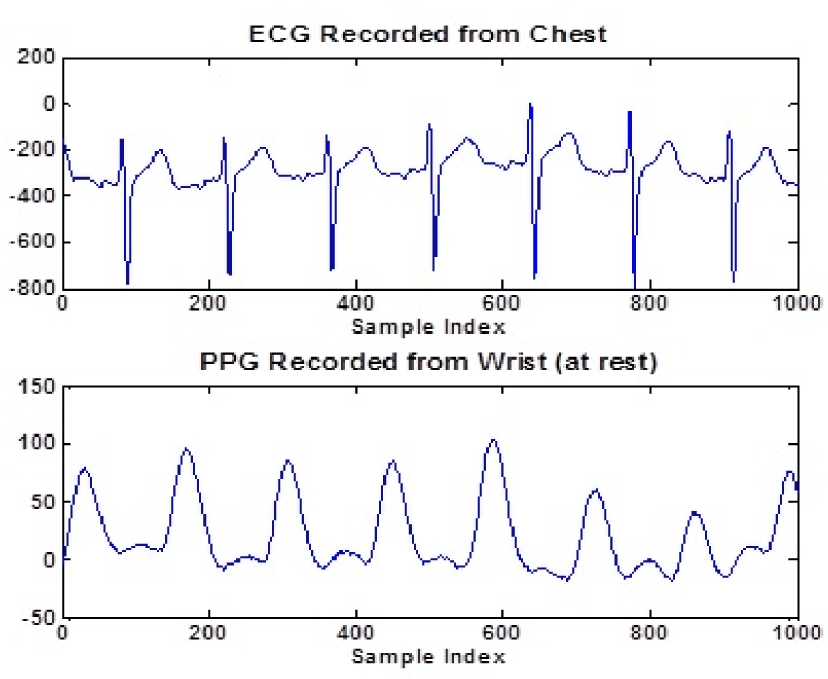
Sample ECG and PPG signal illustrating chest and wrist band calculations at rest and during physical activity

## 3 Self-Supervised Learning Techniques

Self-supervised learning techniques can be valuable for biomedical signal processing, including Electrocardiogram (ECG) and Photoplethysmography (PPG) data analysis. These techniques leverage the abundance of unlabeled data to learn meaningful representations without relying on explicit annotations. Lee et al. [8] introduced a novel self-supervised learning algorithm utilizing ECG delineation, demonstrating its efficacy in classifying arrhythmias. The algorithm significantly improved the performance of deep neural networks (DNNs) across multiple datasets and different levels of labeled data. Additionally, we employed transfer learning to fine-tune the algorithm, further enhancing its capabilities. The pursuit of training CNNs for ECG classification is often hindered by the need for an extensive collection of annotated samples, a costly endeavor. In an ingenious approach, [9] resolved this challenge through the utilization of transfer learning. Initially, CNNs are pretrained on the vast public repository of unprocessed ECG signals. Subsequently, these networks are refined on a limited dataset to specialize in the identification of Atrial Fibrillation, the prevailing cardiac arrhythmia. A new approach called Segment Origin Prediction (SOP) was introduced as a self-supervised pre-training method proposed by Luo et al. [10] aimed at enhancing the model’s ability to classify arrhythmias. By incorporating a data reorganization module, the model learns ECG features by predicting whether two segments belong to the same original signal, eliminating the need for annotations [11]. Additionally, by including a feed-forward layer during the pre-training phase, the model achieves improved performance when utilizing labeled data for arrhythmia classification in subsequent stage [12]. [13] introduces a self-supervised learning approach for precise heart rate estimation. By skillfully approximating a participant’s PPG signal from videos, specifically focusing on well-covered skin regions, their framework triumphantly handles diverse local noises. Additionally, the model enriches an extracted rPPG signal from a facial landmark with neighboring facial landmarks’ rPPG signals, expertly aligning their distinctive personalized patterns.

Jangwon et al. [14] utilized EfficientNet-B3, a powerful deep learning model, as the foundation for their research. They employed self-supervised learning in their pre-training process, specifically using label masking, to handle data from multiple sources. Their goal was to identify clinical diagnoses from various types of ECG recordings, ranging from 12-lead to reduced-lead formats such as 6-lead, 4-lead, 3-lead, and 2-lead.

### 3.1 Contrastive Learning

Contrastive learning is primarily an unsupervised learning technique, where the labels or annotations are not required during the training process. However, in the context of ECG (Electrocardiogram) and PPG (Photoplethysmogram) signal processing, contrastive learning can be combined with a supervised approach to enhance the performance of the models.

#### 3.1.1 Application of contrastive learning to ECG and PPG signals

Contrastive learning has shown great potential in the application of electrocardiogram (ECG) and photoplethysmogram (PPG) signals, providing several benefits in their analysis. One application is in unsupervised representation learning, where contrastive learning can extract meaningful features from large amounts of unlabeled ECG and PPG data. By contrasting similar and dissimilar signal segments, contrastive learning can capture underlying patterns and structure, enabling the model to learn representations that reflect the intrinsic characteristics of the signals. These learned representations can then be utilized for various downstream tasks such as arrhythmia detection, heart rate variability analysis, or blood pressure estimation.

Furthermore, contrastive learning can be employed for domain adaptation and transfer learning in ECG and PPG analysis. By pre-training a model on a large dataset from a source domain, such as a different hospital or population, and then fine-tuning it on a smaller labeled dataset from the target domain, the model can adapt and generalize well to the specific characteristics of the target domain. This is particularly useful in scenarios where labeled data in the target domain is limited or expensive to obtain. Contrastive learning allows the model to leverage the abundant unlabeled data from the source domain to learn robust and transferable representations, improving the performance and generalization capability on the target domain.

However, applying contrastive learning to ECG and PPG signals also presents challenges. One challenge is the selection of appropriate positive and negative pairs for the contrastive loss calculation. In the case of ECG and PPG signals, determining similar and dissimilar segments can be non-trivial due to variations caused by different physiological conditions, noise, artifacts, or measurement devices. Careful consideration is required to design effective strategies for constructing the positive and negative pairs that capture the relevant similarities and differences in the signals. Additionally, the computational complexity of contrastive learning can be demanding, especially when dealing with long ECG or PPG sequences. Efficient sampling methods and optimization techniques need to be employed to make the training process feasible and scalable.

In summary, contrastive learning offers valuable applications in ECG and PPG signal analysis, including unsupervised representation learning and domain adaptation. By leveraging unlabeled data and learning meaningful representations, contrastive learning can enhance the performance of various tasks in ECG and PPG analysis. However, careful consideration of pair construction and computational efficiency is essential to address the challenges specific to these signal types. A novel approach was introduced by John et al. [15] for determining the heart rate of individuals using facial video. Instead of relying on existing blood pressure data, this method focuses on extracting the photoplethysmography (PPG) signal present in facial expressions through self-supervised contrastive learning. To enhance the accuracy, a supervised heart rate estimator is trained to closely align with the actual heart rate measurements. Ye et al. [16] introduces a new technique called Discriminative Clustering Enhanced Adversarial Domain Adaptation (DC-ADA) for detecting mental stress across different individuals using ECG data. This method enhances the training process of existing bi-classifier adversarial UDA methods by incorporating a self-supervised loss on the target data. The proposed approach involves obtaining pseudo labels for the target data through a clustering-based strategy. These labels are then utilized as supervision to guide the classification of ambiguous target data into the appropriate stress levels.

Retraining models for new users in healthcare applications often necessitates the collection of a significant amount of labeled data, which can be both challenging and expensive. This is particularly true for applications like atrial fibrillation detection. To address the scarcity of labeled data, unsupervised and self-supervised techniques have emerged as promising solutions. Fonseca et al. [17] proposes the utilization of contrastive learning to enhance the performance of a CNN that classifies atrial fibrillation in scenarios with limited labeled data, small models, and noisy data. The effectiveness of this strategy was evaluated using the largest publicly available ECG dataset, and the study presents results in terms of the F1-score for various proportions of unlabeled-labeled data and different model sizes. The findings indicate that the proposed strategy surpasses the baseline approach, achieving up to a 30% improvement in the 10-fold mean F1-score compared to a 5.8% enhancement in AUC, which is the current state-of-the-art performance metric.

Remote Photoplethysmography (rPPG) systems offer a non-contact, costeffective, and widely available means of regularly monitoring heart rate (HR) [18]. They achieve this by analyzing the scattered light reflected from changes in blood volume in human skin tissues (known as PPG). However, these systems have not been widely adopted due to the absence of a standardized approach for estimating HR from skin videos in various practical situations. Conventional supervised methods necessitate a substantial number of accurately synchronized annotations between video and rPPG signals, which significantly hinder the development of comprehensive end-to-end rPPG models. Hasan et al. [19] introduces Self-rPPG, a technique that directly learns the optical and physiological mechanisms of rPPG from unlabeled videos without requiring synchronized rPPG signal annotations. The authors devised a self-supervised contrastive learning-based pretraining strategy to learn the representation of the inherent frequency and phase of diffusion signals, as well as the temporal coherence of video frames, using unlabeled sequences collected from multiple public datasets. They conducted experiments to determine the optimal contrastive learning schemes (loss functions and sampling strategy) and to evaluate the significance of the features learned by Self-rPPG. The results showed that the self-supervised representations can effectively encode the frequency and phase of diffusion signals, while also demonstrating resilience against temporal distortion. The effectiveness of different augmentations for contrastive self-supervised learning of electrocardiogram (ECG) signals and the optimal parameters are thoroughly examined in this study [20]. The baseline approach comprises two primary components: contrastive learning and the downstream task. Initially, an encoder is trained using various augmentations to extract ECG signal representations that are generalizable. Subsequently, the encoder is frozen, and a few linear layers are fine-tuned using different quantities of labeled data for arrhythmia detection in downstream tasks. [21] introduced ALPINE, a captivating approach named A noveL rPPG technique for Improving remote heart rate estimatioN using contrastive lEarning. ALPINE incorporates the power of contrastive learning to overcome limited labeled data, fostering data diversity for enhanced network generalization. Moreover, they introduce an innovative hybrid loss, encompassing contrastive loss, signal-to-noise ratio (SNR) loss, and data fidelity loss, further enhancing their methodology.

[21] introduces a remarkable unsupervised learning framework for signal regression, eliminating the reliance on labeled video data. By embracing the essence of periodicity and finite bandwidth, this approach unveils the blood volume pulse from unlabelled videos. The study concludes that promoting sparse power spectra within the realm of normal physiological frequencies, coupled with variance across batches of power spectra, proves ample for learning the visual attributes of periodic signals. [23] introduced a captivating Intra-Inter Subject Self-Supervised Learning (ISL) model tailored for multivariate cardiac signals. This model skillfully incorporates medical expertise to effectively glean insights from differences within and between subjects. For intra-subject self-supervision, the ISL model harnesses a channel-wise attentional CNN-RNN encoder to extract heartbeat-level features from each individual. Additionally, a stationarity test module captures temporal dependencies among heartbeats. In the realm of inter-subject self-supervision, the model employs a series of data augmentations inspired by the unique clinical attributes of cardiac signals. By employing contrastive learning, it acquires distinct representations for different patient types. [24] presents a captivating framework called CROCS, which employs supervised contrastive learning. Here, cardiac signal representations linked to specific patient attributes, such as disease class, sex, and age, are drawn towards learnable embeddings known as clinical prototypes. These prototypes serve a dual purpose: facilitating the clustering and retrieval of unlabeled cardiac signals based on various patient attributes. [25] presents an exquisite, all-encompassing framework for semi-supervised learning across domains, incorporating the elegance of contrastive learning and the prowess of adversarial training strategies. [26] introduced an exquisite unsupervised contrastive learning framework. This framework ingeniously incorporates a fresh contrastive loss, harnessing the power of a data augmentation scheme that gracefully combines two data samples to create novel instances. The central objective of this framework lies in predicting the mixing component, which gracefully serves as soft targets within the loss function. [27] present an exquisite contrastive learning framework that harnesses metadata to curate positive and negative pairs during the training of unlabeled data. Specifically, the contrastive learning model ingeniously integrates metadata from clinical records, such as age, sex, weight, and sound location, to establish a profound connection between lung and heart sound recordings on a patient level, effectively enhancing the overall context.

Proposing a novel technique for remote photoplethysmography (rPPG), [28] introduced an innovative approach that eliminates the need for costly physiological training data. Their method leverages self-supervised training, combining contrastive learning with a subtle prior on the frequency and smoothness of the target signal. By integrating a learned saliency resampling module, they demonstrated the ability to reduce reliance on manually engineered features while offering insights into the model’s behavior and potential limitations.

[29]introduces an exquisite deep learning framework that effortlessly captures the essence of physiological waveforms. By employing convolutional neural networks (CNNs), it distinguishes genuine arrhythmia alarms from false ones. Through the ingenious utilization of Contrastive Learning, the framework minimizes a binary cross entropy classification loss while incorporating a remarkable similarity loss derived from pairwise waveform segment comparisons. Moreover, to enhance the models, they integrate learned embeddings from a rule-based approach, effectively harnessing prior domain knowledge specific to each alarm type.

[30] present an elegant solution called the Contrastive Accelerometer–Gyroscope Embedding (CAGE) model. This innovative approach utilizes a two-stream convolutional neural network (CNN) to process the accelerometer and gyroscope signals independently. By fusing modality-specific features at the feature-level, the model effectively tackles recognition tasks. Additionally, they introduce a self-supervised learning (SSL) task, where the accelerometer and gyroscope embeddings from the same activity instance are paired. In their work, [31] introduced the Dense Lead Contrast (DLC) approach, which utilizes contrastive learning on multilead ECGs. By employing a multibranch network (MBN), DLC fosters intralead and interlead invariance, resulting in a comprehensive global representation through a joint loss that combines intralead and interlead contrastive losses.

#### 3.1.2 Benefits and challenges of contrastive learning in biomedical signal analysis

Contrastive learning has gained attention as a promising technique in biomedical signal analysis, offering several benefits in this domain. One key advantage is its ability to leverage large amounts of unlabeled data to learn meaningful representations. Biomedical signal datasets are often vast and unlabeled, making contrastive learning an attractive approach for unsupervised representation learning. By contrasting positive pairs (similar samples) and negative pairs (dissimilar samples), contrastive learning can capture the underlying structure and patterns in the signals, facilitating subsequent downstream tasks such as classification or anomaly detection. Moreover, contrastive learning enables transfer learning, where models pre-trained on a large dataset can be finetuned on smaller labeled datasets, allowing for efficient utilization of limited labeled biomedical signal data.

However, contrastive learning also presents certain challenges in the context of biomedical signal analysis. One significant challenge is the choice of appropriate similarity metrics or contrastive loss functions. Selecting the right metrics that capture the relevant similarities between signals is crucial for the success of contrastive learning. Additionally, contrastive learning may face difficulties in handling highly imbalanced datasets or rare events in biomedical signals. This can lead to suboptimal representation learning, as the model might prioritize the majority class, potentially overlooking critical minority patterns. Another challenge is the computational complexity of contrastive learning, which often requires significant computational resources and time, particularly when dealing with large-scale biomedical signal datasets. Careful optimization strategies and efficient implementation are necessary to mitigate these challenges and ensure the practicality and effectiveness of contrastive learning in biomedical signal analysis.

### 3.2 Generative Models

Generative models, such as Variational Autoencoders (VAEs) or Generative Adversarial Networks (GANs), can also be employed for self-supervised learning. These models learn to generate realistic synthetic ECG or PPG signals by modeling the underlying data distribution. By training such models on large amounts of unlabeled data, they can capture the inherent structure and patterns of the signals. The trained generative models can then be used for data augmentation, anomaly detection, or as a source of synthetic data for downstream tasks. In their work, [32] introduce CardioGAN, an elegant adversarial framework that transforms PPG signals into exquisite ECG waveforms. This innovative model harnesses an attention-based generator, gracefully capturing intricate details, while employing dual discriminators to safeguard the fidelity of the generated data across both time and frequency domains.

#### 3.2.1 AutoEncoders

Autoencoders are neural network architectures that learn to encode input data into a low-dimensional latent space and then decode it back to the original input. By training an autoencoder on a large amount of unlabeled ECG or PPG data, the model can learn to capture the salient features and patterns within the signals. Once trained, the encoder part of the network can be used to extract meaningful representations that can be employed for downstream tasks like anomaly detection or classification. Daniel et al. [33] presented a novel approach to self-supervised learning specifically for 12-lead electrocardiograms (ECGs). In their method, they introduced a task to mask certain segments of the input signals across all channels and aimed to predict the original values. To implement this, they employed a U-ResNet model consisting of an encoder-decoder structure. They evaluated the model’s performance by initially pretraining it on the CODE dataset using self-supervised learning and subsequently fine-tuning it on the PTB-XL and CPSC benchmarks to leverage the acquired feature.

Dezaki et al. [34] The Echo-Rhythm Net method, proposed in [], introduces an automated approach for detecting atrial fibrillation (AF) by relying solely on echocardiogram imagery (echo), eliminating the requirement for an electrocardiogram (ECG). The framework comprises three primary elements: an encoder, trained through a self-supervised technique, a layer for temporal self-similarity matrix, and a supervised detector that is trained using labels of cardiac rhythm provided by sonographers.

In their study, Yang et al. [35] present a model for learning representations of ECG signals based on a masked autoencoder. The proposed approach involves applying a high masking ratio to the original ECG signal and employing an autoencoder to reconstruct it. The model incorporates both local and global ECG features by utilizing multi-scale convolution for extracting local features and employing a transformer for capturing global features. The training process involves pre-training the model on ECG datasets and subsequently fine-tuning it for each specific ECG classification task.

Catering to the core challenge in diagnostics, [36] addresses the harmonious integration of diverse modalities, forging a unified portrayal of the physiological state. Their ingenious solution lies in a cross-modal autoencoder framework, melding distinct data modalities to create a comprehensive depiction of the cardiovascular condition. Remarkably, this framework fuses cardiac magnetic resonance images (MRIs) that capture structural details with electrocardiograms (ECGs) that convey myoelectric insights, yielding cross-modal representations of exceptional depth and breadth

#### 3.2.2 Use of autoencoders for ECG and PPG signal representation learning

Autoencoders have proven to be valuable tools for representation learning in various domains, including the analysis of electrocardiogram (ECG) and photoplethysmogram (PPG) signals. ECG and PPG signals contain valuable information about the cardiovascular system and can be used for various clinical applications, such as arrhythmia detection, heart rate variability analysis, and blood pressure estimation.

Autoencoders are neural network architectures that are designed to reconstruct their input data. They consist of an encoder network that compresses the input into a lower-dimensional representation, called the latent space, and a decoder network that attempts to reconstruct the original input from the latent representation. By training the autoencoder to minimize the reconstruction error, the model learns to extract meaningful features and representations from the input data.

### 3.3 Variational autoencoders

Variational Autoencoders (VAEs) can indeed be used for self-supervised learning with biomedical signals such as ECG (electrocardiogram) and PPG (photoplethysmogram). VAEs are a type of generative model that can learn the underlying latent representation of the data by training on unlabeled samples. This latent representation can then be used for various downstream tasks, including classification, anomaly detection, and data generation

#### 3.3.1 Introduction to VAEs

Variational Autoencoders (VAEs) are a type of generative model that can be used for self-supervised learning. Self-supervised learning is an approach to training machine learning models where the labels or annotations for the training data are automatically generated from the data itself, rather than being provided by human annotators.

In the context of VAEs, self-supervised learning typically involves training the model to reconstruct the input data, or to generate new samples that are similar to the input data. The basic idea is to use the unsupervised training process of the VAE to learn a latent representation of the data that captures its salient features, without relying on any external annotations.

#### 3.3.2 Application of VAEs for ECG and PPG signal feature extraction

Variational Autoencoders (VAEs) have found applications in various domains, including biomedical signal processing such as electrocardiogram (ECG) and photoplethysmogram (PPG) signal analysis. VAEs can be employed for feature extraction from ECG and PPG signals, which are vital in diagnosing cardiovascular diseases and monitoring physiological conditions. By leveraging the unsupervised learning capabilities of VAEs, the encoder-decoder architecture can learn to capture the underlying patterns and distinctive characteristics of these signals in a latent space representation. This latent space can then be utilized to extract relevant features such as heart rate variability, rhythm abnormalities, or pulse characteristics. The reconstructed signals obtained from the decoder can be compared with the original input signals, enabling the evaluation of reconstruction loss and the accuracy of the learned representation. By applying VAEs for ECG and PPG signal feature extraction, researchers and clinicians can potentially improve the accuracy of disease detection, monitoring, and risk assessment, leading to more effective healthcare interventions.Huang et al. [37] introduced a captivating and ingenious self-supervised pretraining method. Unveiling an adept encoder that gracefully acquires spatiotemporal representations by skillfully reconstructing 12-lead ECG signals. These signals, artfully masked in both time and lead dimensions, reveal the true brilliance of this novel approach.

#### 3.3.3 Discussing the strengths and weaknesses of VAEs in biomedical signal analysis

Variational Autoencoders (VAEs) have emerged as powerful tools in biomedical signal analysis, offering several strengths that make them valuable in this field. One of their main strengths lies in their ability to capture complex, highdimensional patterns in biomedical signals, such as electrocardiograms (ECGs) or electroencephalograms (EEGs). VAEs can learn a latent representation of the signals, enabling efficient data compression and reconstruction. This can be particularly useful in cases where data storage or transmission constraints are present. Furthermore, VAEs provide a probabilistic framework, enabling uncertainty estimation and generating synthetic samples, which can be valuable for data augmentation and generating simulated datasets for training.

However, VAEs also possess certain weaknesses that should be considered in biomedical signal analysis. One limitation is the difficulty in guaranteeing the preservation of the fine details and temporal dynamics of the original signals during the compression and reconstruction process. The reconstruction quality might be compromised, especially in cases where the input signals contain subtle but clinically relevant information. Additionally, VAEs heavily rely on the assumption of independence between the elements of the latent space, which may not always hold true in complex biomedical signal analysis. Another challenge is the selection of appropriate hyperparameters, such as the dimensionality of the latent space, which can significantly impact the performance and interpretability of VAE models. Therefore, careful tuning and validation are essential to ensure optimal results when using VAEs in biomedical signal analysis.

### 3.4 Other self-supervised learning techniques

#### 3.4.1 Predictive coding

Contrastive predictive coding (CPC) is a self-supervised learning framework that aims to capture the underlying structure of the data. It involves training a model to predict future segments of the signal given previous segments. By maximizing the agreement between predictions and the actual future segments, the model learns to capture relevant features and patterns in the data. With the adept use of GNN pre-training techniques, the authors [38] magnify the potential of Graphene by retraining it with a graph attention architecture. This innovative approach triumphs over rival methods in the realms of pathway gene recovery, disease gene reprioritization, and comorbidity prediction, showcasing unparalleled performance.

#### 3.4.2 Temporal alignment

Temporal alignment models are designed to learn temporal dependencies within a sequence of data. In the case of ECG and PPG, these models can learn to align and compare different parts of the signals to identify meaningful patterns. By training these models on unlabeled data, they can learn to capture temporal relationships between different signal segments, which can be beneficial for tasks like heartbeat segmentation or event detection. In order to effectively utilize time-series signals like physiological signals, it is crucial that the representations employed capture pertinent information from the entire signal. In a study conducted by Juan et al. [39], a Transformer-based model was proposed for the analysis of electrocardiograms (ECG) in the context of emotion recognition. The attention mechanisms of the Transformer were utilized to construct contextualized representations of the signal, placing greater emphasis on relevant portions. These representations were subsequently processed using a fully-connected network to predict emotions. To address the limited availability of emotional labels in datasets, the researchers employed self-supervised learning. They collected multiple ECG datasets lacking emotion labels and employed them for pre-training their model, which was then fine-tuned using the AMIGOS dataset to perform emotion recognition. [40] propose a novel approach called Mixed Supervised Contrastive Loss (MSCL) for MTS representation learning. This approach combines self-supervised, intra-class, and inter-class supervised contrastive learning to effectively utilize labels. Building upon MSCL, they introduce a new framework named MIxed supervised COntrastive learning for MTS classification (MICOS), which leverages spatial and temporal channels to extract complex spatio-temporal features from MTS data. [41] introduce TimeCLR, a framework for univariate time series representation. It combines DTW and InceptionTime to create a feature extractor with strong feature extraction capabilities. The approach involves DTW data augmentation to generate targeted phase shift and amplitude change phenomena while preserving time series structure and features. By leveraging the advantages of both DTW data augmentation and InceptionTime, TimeCLR extends SimCLR and finds successful applications in the time series domain. A multimodality ECG classification approach using SSL was proposed by [20]. The approach follows the SSL learning paradigm and comprises two modules: one for the pre-stream task and another for the downstream task. In the SSL-pre-stream task, they applied self-knowledge distillation (KD) techniques without using labeled data, considering various transformations in both the time and frequency domains. For the downstream task, which utilizes labeled data, they introduced a gate fusion mechanism to merge information from different modalities. [42] introduced the T-S reverse detection, an elegant and powerful self-supervised method to acquire ECG representations. Harnessing the temporal and spatial qualities of ECG signals, this approach ingeniously employed horizontal, vertical, and combined temporal-spatial reversals. By categorizing four signal types, including the original, it facilitated learning.

#### 3.4.3 Multitask learning

Multitask learning involves training a model to perform multiple related tasks simultaneously. In the context of ECG and PPG analysis, you can define multiple tasks such as heart rate estimation, anomaly detection, or signal denoising. By jointly training the model on multiple tasks, it can learn to extract useful representations that are relevant across different tasks. An elegant multitask learning framework, CO-TASK, was introduced by [43]. It enhances the performance of multi-task learning by creating auxiliary tasks through the harmonious fusion of task labels. CO-TASK ingeniously generates these auxiliary tasks without requiring extra labeling, exhibits resilience towards label noise, and seamlessly integrates with diverse multi-task learning techniques.

## 4 Applications of Self-Supervised Learning in ECG and PPG Analysis

Self-supervised learning is a powerful technique in machine learning where a model learns from unlabeled data by creating surrogate labels from the data itself, rather than relying on human-labeled datasets. This approach has shown great potential in various domains, including ECG (Electrocardiogram) and PPG (Photoplethysmogram) analysis [44] introduced a groundbreaking convolutional architecture, which capitalizes on casual dilated convolutions and residual connections to extend effective memory and achieve outstanding performance. By leveraging raw Electrocardiograph (ECG) and Photoplethysmograph (PPG) signals directly, without the need for hand-crafted feature extraction like conventional methods, they unlock the true potential of deep learning. This approach enhances the extraction of intrinsic features (deep features) and improves long-term robustness [45]. The study suggests a deep convolutional neural network with attention mechanisms for ECG emotion recognition. To account for individuality differences, an enhanced Convolutional Block Attention Module (CBAM) is integrated into the network. CBAM’s channel attention assigns weights to ECG features from various channels, while spatial attention provides weighted representations of ECG features within each channel [46]. [47] employ a self-supervised deep multi-task learning approach for emotion recognition using electrocardiogram (ECG) data. They first learn ECG representations through a signal transformation recognition network using unlabeled data. Then, they transfer the network’s weights to an emotion recognition network, freezing the convolutional layers and training the dense layers with labeled ECG data. [48] presented an automated framework for inferring personalized 4D surface meshes of cardiac structures from 2D echocardiography videos. This step is crucial for precise personalized simulation and automated assessment of cardiac chamber morphology and function. The method is trained using unpaired echocardiography and heart mesh videos, employing a self-supervised approach to find the mapping between these visual domains. The study [49] emphasizes self-supervised techniques and models in healthcare, exploring their benefits and constraints in tasks involving electronic health records, medical images, bioelectrical signals, genes, and proteins. Additionally, it explores potential applications of selfsupervised learning with multimodal datasets and the challenges of acquiring unbiased data for training. Overall, self-supervised learning holds the promise of speeding up the advancement of medical artificial intelligence.

### 4.1 ECG analysis

Analyzing electrocardiogram (ECG) signals using self-supervised learning is an interesting and promising approach. It is an essential task in cardiology to diagnose various heart-related conditions and has gained popularity in various fields due to its ability to leverage large amounts of unlabeled data and extract meaningful representations. In their work, [50] introduced an emotion recognition system based on ECG using self-supervised learning. The system comprises two key networks: a signal transformation recognition network and an emotion recognition network. Initially, the former network is trained on unlabeled data to detect predefined signal transformations through self-supervised learning. Subsequently, the convolutional layer weights of this network are transferred to the emotion recognition network, which is further trained with two dense layers to classify arousal and valence scores. In their work, [51] introduce Echo-SyncNet, a self-supervised learning approach for synchronizing 2D echo series without human supervision or external inputs. The framework utilizes two types of supervisory signals: spatiotemporal patterns within a cine (intra-view self-supervision) and interdependencies between multiple cines (inter-view self-supervision). By leveraging these signals, the model learns a feature-rich and low-dimensional embedding space to achieve temporal synchronization of multiple echo cines

#### 4.1.1 Rhythm analysis and abnormality detection

Li et al. [52] introduces DrCubic an innovative method for categorizing heart irregularities by utilizing condensed-lead ECG recordings. Their approach includes integrating peak detection as a self-supervised auxiliary task and constructing the model around SE-ResNet. They also incorporate models with varying input lengths and sampling rates to ensure optimal performance within a single source and across multiple sources simultaneously.

#### 4.1.2 QRS complex detection and delineation

QRS complex detection and delineation is a crucial task in the field of electrocardiogram (ECG) and photoplethysmogram (PPG) analysis for accurately identifying cardiac events. Self-supervised learning has emerged as a promising approach to tackle this challenge effectively. By utilizing large volumes of unlabeled ECG and PPG data, self-supervised learning algorithms can autonomously learn relevant representations and patterns within the signals. Through this process, the model can develop a robust understanding of QRS complexes, enabling it to detect and delineate these critical features accurately. This innovative approach eliminates the need for costly and time-consuming manual annotation, making it highly scalable and adaptable to different datasets and scenarios. The potential of self-supervised learning in QRS complex detection and delineation holds tremendous promise for advancing the field of cardiovascular research and enhancing the accuracy of cardiac monitoring and diagnosis in real-world healthcare applications. According to [53], the proposed method involves framing the signal based on the detected QRS complex (R peaks). As a result, consecutive frames of the signal exhibit high similarity, reducing redundancy and increasing sparsity. To enhance detection performance, frames indicating cardiovascular disease symptoms are transmitted uncompressed. The paper [54] introduces a system that utilizes smart speakers to capture heartbeats without physical contact. By employing algorithms that convert the smart speaker into a short-range active sonar system, they measure heart rate and inter-beat intervals (R-R intervals) for regular and irregular rhythms. The smart speaker emits inaudible 18–22 kHz sound and detects echoes from the human body, which encode sub-mm displacements caused by heartbeats. A new architecture LinkNet++ [55] is introduced to efficiently and automatically extract fECG from abdominal mECG. It utilizes a feature-addition approach, combining deep and shallow levels with residual blocks, addressing the shortcomings of U-Net and UNet++ models. The model’s performance was assessed for fECG signal reconstruction and fetal QRS (fQRS) detection.

#### 4.1.3 Heart rate variability (HRV) analysis

Heart rate variability (HRV) analysis is a physiological measurement that assesses the variations in time intervals between successive heartbeats. It is a non-invasive method widely used to evaluate the autonomic nervous system (ANS) activity, specifically the balance between the sympathetic and parasympathetic branches. HRV is considered an important indicator of the body’s ability to adapt to various stressors and reflects the overall health and well-being of an individual. HRV analysis finds applications in various fields, including cardiology, stress management, sports performance, and general health assessment. It can help identify autonomic imbalances, assess cardiovascular risk, evaluate the effectiveness of interventions, and provide insights into an individual’s response to stress or training. However, it’s essential to interpret HRV data within the context of other clinical and physiological parameters to draw meaningful conclusions.

1. [56] introduces a transductive meta-learner that leverages unlabeled samples during testing for self-supervised weight adjustment, enabling swift adaptation to distributional changes in Remote heart rate estimation using remote photoplethysmography (rPPG).

A new self-supervised representation learning framework called ”Contrastive Heartbeats (CT-HB)” [57] is proposed to develop general and robust electrocardiogram representations for efficient training on different downstream tasks. This framework uses a unique heartbeat sampling method to create positive and negative pairs of heartbeats for contrastive learning, leveraging the meaningful patterns in electrocardiogram signals. Through CT-HB, the self-supervised learning model acquires personalized heartbeat representations that reflect the patient’s specific cardiology contex

#### 4.1.4 Arrhythmia classification and prediction

Arrhythmia refers to an abnormal heart rhythm that deviates from the normal sinus rhythm. It can manifest as irregular heartbeats (too fast, too slow, or irregular pattern) and may result from various factors such as heart disease, electrolyte imbalances, drug use, or genetic conditions. Arrhythmia classification and prediction involve identifying and categorizing different types of arrhythmias and assessing the likelihood of their occurrence. This process is crucial for the early detection, treatment, and management of cardiac arrhythmias. ECG is the primary tool for diagnosing arrhythmias. It records the electrical activity of the heart over time, representing it as a graph. Cardiologists and algorithms can analyze the ECG signals to identify various types of arrhythmias based on specific patterns in the ECG waveform. [58] endeavors to develop a precise screening method for arrhythmia (ARR), aiding physicians in diagnosing heart diseases more effectively. The proposed approach introduces a multi-domain feature extraction technique and a hierarchical extreme learning machine (H-ELM) network to predict fetal ARR. Initially, the multidomain feature extraction technology captures rich, high-dimensional features that represent the original signal. Next, the sensitive features are identified through neighborhood component analysis (NCA) from the high-dimensional vectors. These sensitive features are then fed into a stacked extreme learning machine sparse autoencoder (ELM-SAE), which employs a layer-by-layer unsupervised learning process to extract high-level fusion features. Finally, an original ELM is integrated into the ELM-SAE network for accurate fetal ARR prediction. The study[59] proposes an S-shaped reconstruction method to identify arrhythmia. It involves data preprocessing, denoising the original ECG data, removing baseline drift, extracting heartbeats, and balancing data using a synthetic minority oversampling technique algorithm. The method converts the one-dimensional heartbeat series into a 2-D matrix to analyze relationships between distant points in an ECG series. Finally, a 2-D 19-layer SE-ResNet is employed to categorize heartbeats into normal, supraventricular ectopic, ventricular ectopic, fusion, and unknown beats. [60] introduces supervised contrastive learning (sCL), a model that utilizes labeled data to bring instances of the same class closer while pushing different classes apart, mitigating the risk of false negatives for arrhythmia classification. [61] introduce CLECG, a new instance-level contrastive learning method for ECG signals, aiming to extract valuable information from unlabeled data. CLECG encourages similar representations for augmented views of the same signal (positive samples) and enhances the distance between representations of augmented views from different signals (negative samples) during pre-training. The MAE-based MaeFE network, proposed in [62], includes three customized masking modes: masked time autoencoder (MTAE), masked lead autoencoder (MLAE), and masked lead and time autoencoder (MLTAE). MTAE and MLAE focus on temporal and spatial features, respectively, while MLTAE combines both. During pretraining, ECG signals are divided into patches and partially masked for encoder-token transfer and decoder reconstruction. In downstream tasks, the pretrained encoder serves as an arrhythmia classifier for the downstream dataset.

### 4.2 PPG Analysis

Analyzing PPG signals using self-supervised learning is an interesting approach that involves leveraging the data itself to create labels for training without human annotations. Self-supervised learning is a subset of unsupervised learning, where the model learns from the input data indirectly. In the case of PPG analysis, self-supervised learning can be used to extract meaningful representations and features from the raw PPG signal.

#### 4.2.1 Blood pressure estimation

Blood pressure estimation using electrocardiogram (ECG) and photoplethysmogram (PPG) through self-supervised learning represents a groundbreaking approach in the field of healthcare technology. By leveraging self-supervised learning techniques, the system can learn from a vast amount of unlabeled data, which substantially reduces the reliance on costly and time-consuming annotated datasets. This novel method harnesses the synergistic information present in both ECG and PPG signals, capturing intricate cardiovascular dynamics for precise blood pressure estimation. Through unsupervised feature learning, the system can extract relevant patterns and correlations, leading to more accurate and robust predictions. By fusing the power of ECG and PPG data with self-supervised learning, this cutting-edge technology has the potential to revolutionize blood pressure monitoring, enabling non-invasive, continuous, and real-time assessment of cardiovascular health in a cost-effective and user-friendly manner.

#### 4.2.2 Pulse rate variability (PRV) analysis

Pulse Rate Variability (PRV) analysis, using Electrocardiogram (ECG) and Photoplethysmogram (PPG) data, has witnessed significant advancements through the implementation of self-supervised learning techniques. PRV refers to the variation in the time interval between successive heartbeats and provides valuable insights into the autonomic nervous system’s functioning and overall cardiovascular health. Self-supervised learning is an emerging approach that leverages unlabeled data to train models without requiring manual annotations. In the context of PRV analysis, this technique enables the extraction of meaningful patterns and features from large-scale ECG and PPG datasets. By learning to predict and reconstruct underlying temporal structures within the data, self-supervised models can subsequently be fine-tuned for various cardiovascular tasks, such as arrhythmia detection, stress assessment, and heart rate monitoring. This innovative approach holds great promise for enhancing the accuracy and efficiency of PRV analysis, ultimately contributing to improved diagnosis and management of cardiovascular conditions.

#### 4.2.3 Oxygen saturation (SpO2) estimation

Oxygen saturation (SpO2) estimation is a critical parameter used to monitor a patient’s respiratory and cardiovascular health. Traditional methods for SpO2 estimation often rely on specialized medical equipment, such as pulse oximeters, which can be cumbersome and may not be readily available in all settings. However, recent advancements in self-supervised learning techniques have shown promise in accurately estimating SpO2 using simpler and more ubiquitous sensors, such as electrocardiography (ECG) and photoplethysmography (PPG) sensors. Self-supervised learning leverages the vast amount of unlabeled data available from these sensors to learn robust and generalized representations. By capitalizing on the intrinsic relationships between ECG and PPG signals and SpO2 levels, the self-supervised learning model can effectively estimate oxygen saturation levels, paving the way for more accessible and cost-effective healthcare monitoring solutions in various environments. This innovation has the potential to revolutionize remote patient monitoring, home healthcare, and resource-limited medical settings, ultimately improving patient outcomes and healthcare accessibility.

#### 4.2.4 Disease detection and monitoring (e.g, hypertension, sleep apnea)

Disease detection and monitoring using electrocardiogram (ECG) and photoplethysmogram (PPG) data have become increasingly important in modern healthcare. These two non-invasive techniques provide valuable information about a person’s cardiovascular health and can be used to detect and monitor various diseases, including hypertension and sleep apnea.

##### Hypertension Detection and Monitoring

Hypertension is a common condition where the blood pressure in the arteries is consistently elevated. It is a significant risk factor for heart disease, stroke, and other cardiovascular issues. Both ECG and PPG can be used to aid in the detection and monitoring of hypertension. Electrocardiogram records the electrical activity of the heart, which allows medical professionals to assess the heart’s rhythm and identify any abnormalities. In the context of hypertension, ECG can be used to look for signs of left ventricular hypertrophy (LVH), which is an indicator of the heart working harder due to high blood pressure. Photoplethysmogram measures changes in blood volume through the skin. PPG can provide information about the peripheral vascular resistance, which can be associated with hypertension. Additionally, PPG can be used to measure blood pressure indirectly by analyzing the pulse waveform characteristics.

##### Sleep Apnea Detection and Monitoring

Sleep apnea is a sleep disorder characterized by interrupted breathing during sleep, which can lead to reduced oxygen levels and other health issues. ECG and PPG plays a role in diagnosing and monitoring sleep apnea, especially during sleep studies conducted in a clinical setting. During sleep apnea episodes, there are characteristic changes in heart rate and rhythm due to the disruptions in breathing. ECG data can help identify these patterns, especially when synchronized with other sleep monitoring parameters. PPG can be used to assess changes in blood flow and oxygen levels during sleep. When a person experiences sleep apnea events, there may be fluctuations in blood oxygen saturation, which can be detected using PPG-based pulse oximeters.

In [63], a self-supervised representation learning (SSRL) approach is introduced for detecting hypopnea events in single-channel electrocardiography (ECG) signals. The model is trained in two phases: first, an encoder learns signal representation from unlabeled data, and then the classifier and encoder are fine-tuned for classification in the second phase.

### 4.3 Performance Evaluation Metrics for self-supervised learning models in biomedical signal analysis

Performance evaluation and comparison metrics are essential when working with electrocardiogram (ECG) and photoplethysmogram (PPG) signals, as they allow researchers and clinicians to assess the accuracy and effectiveness of different algorithms or methods in processing and analyzing these physiological signals. The common metrics used for evaluating and comparing ECG and PPG signals includes:

#### 4.3.1 Signal Quality Metrics

Signal quality metrics are measurements used to assess the performance and reliability of an ECG or PPG signal. These metrics help in determining the effectiveness of signal transmission and reception, identifying potential issues, and optimizing the overall system performance. The specific metrics used varies depending on the signal i.e ECG or PPG application,here are some common ECG and PPG signal quality metrics:

##### Signal-to-Noise Ratio (SNR)

Measures the ratio of signal power to noise power and indicates the quality of the acquired signal.

##### Signal-to-Interference Ratio (SIR)

Measures the ratio of signal power to interference power, which can be useful in noisy environments.

##### Signal-to-Noise and Distortion Ratio (SNDR)

Combines both noise and distortion measurements in the signal, providing a comprehensive quality assessment.

#### 4.3.2 Accuracy Metrics

Accuracy metrics are important in evaluating the performance of algorithms or models used for ECG and PPG signal analysis. These metrics help to measure how well the algorithm can identify various features or abnormalities in the signals.These metrics are used to evaluate the performance of algorithms for tasks, such as detecting specific ECG waveforms e.g. QRS complexes, P waves or identifying abnormalities in PPG signals e.g., detecting heart rate variability, arterial stiffness. Here are the common accuracy metrics used in ECG and PPG signal analysis:

##### Sensitivity (Recall)

Sensitivity measures the proportion of true positive cases correctly identified by the algorithm. In the context of ECG or PPG analysis, it quantifies how well the algorithm detects the presence of certain features or abnormalities in the signal.

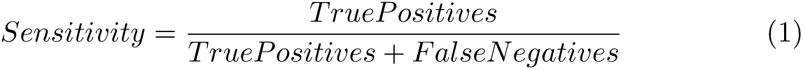

##### Specifity

Specificity measures the proportion of true negative cases correctly identified by the algorithm. It quantifies the ability of the algorithm to correctly rule out the absence of certain features or abnormalities.

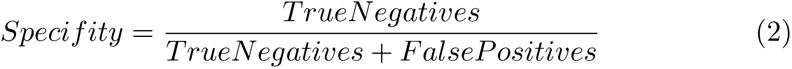

##### Precision

Precision is the proportion of true positive cases out of all the cases predicted as positive by the algorithm. It assesses the algorithm’s accuracy in correctly identifying positive cases.

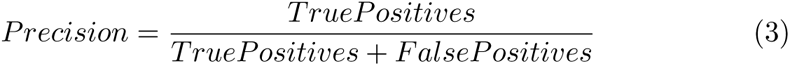

##### Accuracy

Accuracy is the overall proportion of correct predictions made by the algorithm, considering both true positives and true negatives.

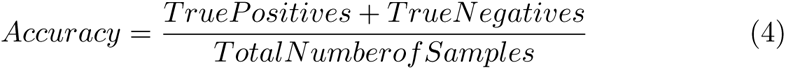

##### F1-Score

The F1 score is the harmonic mean of precision and sensitivity. It provides a balanced measure that considers both false positives and false negatives.

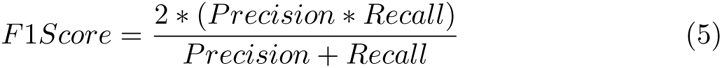

##### Area Under the Curve (AUC)

The AUC is often used to evaluate the performance of binary classification algorithms. It represents the area under the receiver operating characteristic (ROC) curve and measures the algorithm’s ability to distinguish between classes (e.g., normal and abnormal signals)

##### Mean Square Error (MSE)

Measures the average squared difference between the true and estimated signal values.

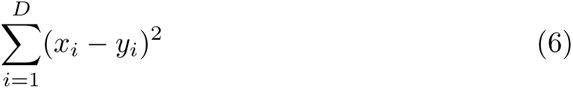

##### Root Mean Square Error (RMSE)

The square root of MSE, providing a measure of the average absolute error between the true and estimated signals.

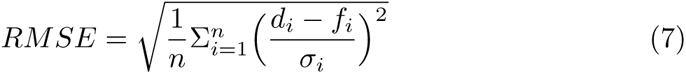

##### Mean Absolute Error (MAE)

Measures the average absolute difference between the true and estimated signal values.

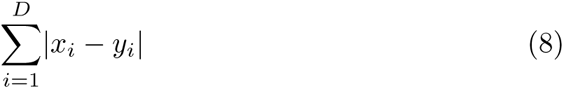

#### 4.3.3 Heart Rate Metrics

Heart rate metrics are essential parameters used in electrocardiogram (ECG) and photoplethysmogram (PPG) analysis to assess the heart’s performance and overall cardiovascular health. Both ECG and PPG are commonly used methods for monitoring heart rate and detecting abnormalities.

##### Heart Rate (HR)

Heart rate represents the number of heartbeats per minute (bpm) and is one of the fundamental metrics for assessing cardiac function. It provides valuable information about the rhythm and overall activity of the heart.

##### Heart Rate Variability (HRV)

HRV is the variation in time intervals between successive heartbeats. It is an essential indicator of the autonomic nervous system’s modulation of heart rate and can provide insights into the heart’s adaptability and response to different physiological and psychological stressors.

##### Maximum Heart Rate (MHR)

The highest heart rate recorded during a specific activity or period is the maximum heart rate. It is often used to set target heart rate zones during exercise or stress testing.

##### Minimum Heart Rate

The lowest heart rate recorded during a specific period is the minimum heart rate. This metric is useful in understanding the resting heart rate or the lowest heart rate achieved during sleep.

##### Heart Rate Recovery (HRR)

HRR is the rate at which the heart rate declines after exercise. It is used as an indicator of cardiovascular fitness and the ability of the autonomic nervous system to recover efficiently after physical exertion.

#### 4.3.4 Time-Domain Metrics

##### R-R Intervals

In ECG analysis, the R-R interval refers to the time duration between successive R-peaks (the highest point of each heartbeat). In PPG analysis, it is referred to as the interbeat interval (IBI), which measures the time between consecutive peaks of the PPG waveform. R-R or IBI intervals are used to calculate heart rate variability (HRV) and assess the autonomic nervous system’s influence on heart rate.

##### Pulse Transit Time (for PPG)

Measures the time it takes for the pulse wave to travel between two arterial sites, often used for blood pressure estimation.

#### 4.3.5 Frequency-Domain Metrics

##### Power Spectral Density (PSD)

Represents the distribution of signal power across different frequency bands, often used in HRV analysis.

#### 4.3.6 Sensitivity and Specificity

If the signals are used for detecting specific events (e.g., arrhythmia, abnormalities), these metrics assess the algorithm’s ability to correctly identify true positive and true negative cases.

#### 4.3.7 Correlation Coefficients

Assess the linear relationship between two signals, such as the cross-correlation coefficient.

#### 4.3.8 Bland-Altman Analysis

Provides a graphical representation of agreement between two methods or signals by plotting the differences against the average of the two measurements.

### 4.4 Performance Comparison with Supervised learning approaches

Supervised learning approaches have been widely used in ECG and PPG analysis, where labeled datasets are essential for training. These methods rely on annotated data to learn patterns and features specific to certain cardiac events or health conditions. By using labeled data, supervised models can achieve high accuracy and precision in detecting abnormal heart rhythms or other cardiac anomalies. However, one of the main challenges with supervised learning lies in the acquisition and labeling of large-scale, high-quality datasets, which can be time-consuming, expensive, and often limited by the availability of expert annotations.

On the other hand, self-supervised learning has emerged as a promising alternative that alleviates the dependence on labeled data. Self-supervised approaches utilize the inherent structure or relationships within the data to create surrogate supervisory signals. For ECG and PPG analysis, this could involve predicting time intervals between different segments of the same signal or predicting a signal segment based on another part of the same signal. By leveraging the data’s intrinsic information, self-supervised methods can efficiently pretrain models on large amounts of unlabeled data. This pretraining stage can then be followed by fine-tuning on a smaller labeled dataset to achieve higher accuracy.

In terms of performance comparison, both approaches have their advantages and limitations. Supervised learning methods typically outperform self-supervised approaches when abundant high-quality labeled data is available. They can learn intricate patterns that may be challenging for self-supervised techniques to capture, leading to better precision and recall in specific diagnostic tasks.

However, in scenarios where labeled data is scarce or expensive to obtain, self-supervised learning shines. These methods offer a cost-effective solution to leverage vast amounts of unlabeled data and extract valuable features, which can then be fine-tuned with limited labeled data to achieve competitive performance. Self-supervised learning has the potential to significantly advance ECG and PPG analysis in resource-constrained settings and enable the development of more robust and generalized model.

### 4.5 Comparative analysis of different self-supervised learning techniques for ECG and PPG signals

Self-supervised learning techniques have shown promising results in various domains, including ECG and PPG signal analysis. In the context of these physiological signals, several self-supervised learning approaches have been compared to assess their efficacy. One widely used method is Contrastive Predictive Coding (CPC), which leverages the contrastive loss to learn meaningful representations. CPC has demonstrated excellent performance in capturing temporal dependencies and underlying patterns within ECG and PPG signals [21]. Another approach is Temporal Alignment [11] (TA) that aligns segments of the same signal, enabling the model to learn informative representations while discarding non-relevant variations. TA has shown promise in handling irregular heartbeats and motion artifacts in PPG signals [19]. Additionally, Generative Pre-trained Transformers (GPT) have been applied to learn hierarchical representations from sequential data, including ECG and PPG signals. GPT-based models have achieved state-of-the-art results in various natural language processing tasks, and their application to physiological signals holds great potential [9]. Overall, these comparative analyses indicate that self-supervised learning techniques can effectively extract meaningful features from ECG and PPG signals, providing valuable insights for cardiovascular health monitoring, disease detection, and personalized healthcare applications. However, the choice of the most suitable technique still depends on the specific dataset characteristics, signal quality, and task requirements.

## 5 Challenges and Future Direction

Self-supervised learning for electrocardiogram (ECG) and photoplethysmogram (PPG) data presents several significant challenges. One of the primary obstacles is the scarcity of large-scale labeled datasets in the medical domain. Both ECG and PPG data require expert annotations for accurate ground truth, making it expensive and time-consuming to acquire sufficient labeled data. Without a substantial amount of labeled data, self-supervised algorithms may struggle to learn robust representations from the limited information available.

Another challenge lies in defining appropriate pretext tasks for self-supervised learning. Designing effective pretext tasks that can capture meaningful representations from raw ECG and PPG signals is critical. These tasks should be carefully crafted to encourage the model to learn relevant features and avoid overfitting to noise or trivial correlations in the data. Finding the right balance between complexity and interpretability of pretext tasks is crucial to ensure the learned representations are clinically meaningful and applicable in real-world scenarios.

Furthermore, the intrinsic complexities of physiological signals, such as ECG and PPG, pose difficulties for self-supervised learning models. These signals are highly dynamic, non-stationary, and subject to various inter and intra-patient variabilities. Extracting informative features from such complex data requires advanced modeling techniques and architectures. Ensuring that the self-supervised models can generalize well across different patients, medical conditions, and measurement conditions is a considerable challenge.

Moreover, addressing the issue of data quality and noise is essential in self-supervised learning for ECG and PPG. Medical signals are often contaminated with artifacts, baseline drift, and other sources of noise, which can mislead the learning process. Developing robust self-supervised algorithms that can effectively handle noisy data and learn resilient representations is critical for achieving reliable and accurate performance.

Lastly, the ethical considerations in medical self-supervised learning cannot be ignored. As these models have the potential to impact patient care and decision-making, ensuring the privacy and security of sensitive patient information is of utmost importance. Striking a balance between model performance and data privacy is a significant challenge in the development and deployment of self-supervised learning models for ECG and PPG data in realworld healthcare settings. Adhering to strict ethical guidelines and regulatory frameworks is crucial to building trustworthy and responsible self-supervised learning systems for medical applications.

In self-supervised learning for ECG (Electrocardiogram) and PPG (Photoplethysmogram) signals, the future holds promising directions to enhance both the accuracy and efficiency of cardiovascular health monitoring. One significant focus is on developing novel pretext tasks and representation learning techniques to leverage the abundant unlabeled data available in real-world scenarios. By designing pretext tasks that exploit the temporal and spectral patterns inherent in ECG and PPG signals, models can learn meaningful representations without requiring extensive annotations.

Moreover, researchers are exploring multi-modal learning approaches, incorporating data from different sources such as accelerometer data, respiratory signals, or patient demographics, to enrich the feature representations and provide a more comprehensive view of an individual’s physiological status. This fusion of information could potentially lead to more robust and personalized predictive models for various cardiovascular conditions.

Another promising direction involves addressing the challenges posed by data imbalance, domain shift, and potential biases in real-world datasets. Techniques such as domain adaptation and transfer learning are being investigated to enable models to generalize better across diverse patient populations and data distributions, making them more applicable in real clinical settings.

Additionally, there is a growing emphasis on the development of self-supervised learning algorithms that are computationally efficient and can be deployed on edge devices or wearable sensors. This can lead to real-time and continuous monitoring of cardiovascular health, providing timely insights and facilitating early detection of anomalies or potential health risks.

Overall, the future of self-supervised learning for ECG and PPG signals appears to be directed towards harnessing unlabeled data effectively, incorporating multiple modalities, addressing data-related challenges, and optimizing for practical implementation in real-world healthcare applications. These advancements hold the potential to revolutionize cardiovascular health monitoring, ultimately leading to improved patient care and preventive healthcare measures

### 5.1 Data Availability

Data availability in self-supervised learning for ECG (electrocardiogram) and PPG (photoplethysmogram) depends on various factors. ECG and PPG data are typically collected from wearable devices, medical sensors, or health monitoring systems. The availability of such data depends on factors like:

#### Data Privacy

ECG and PPG data involve sensitive health information, so access to largescale datasets might be limited due to privacy concerns.

#### Research Institutions

Some research institutions, hospitals, or medical centers may have proprietary datasets, but access to these datasets might require collaborations or specific permissions.

#### Public Datasets

Over time, some publicly available datasets for ECG and PPG might have been released. These datasets can be valuable for research purposes and can help in the development of self-supervised learning methods.

#### Commercial Wearable Devices

Some wearable device companies might have collected substantial amounts of ECG and PPG data from their users, but access to this data might be restricted or require partnership agreements.

### 5.2 Data Quality

The quality of data in self-supervised learning for ECG and PPG is crucial for achieving good performance in the models. Some considerations for data quality include:

#### Noise

ECG and PPG signals can be affected by noise due to various factors such as movement artifacts, sensor malfunctions, or environmental interference. Noise reduction techniques are essential to improve data quality.

#### Labeling Errors

Self-supervised learning often relies on automatically generated labels or pseudo-labels. Errors in labeling can propagate through the learning process and affect model performance.

#### Data Imbalance

Imbalanced datasets can bias the model towards the majority class, leading to suboptimal generalization.

#### Data Preprocessing

Proper preprocessing techniques are necessary to remove artifacts, baseline wander, and other anomalies from the ECG and PPG signals.

### 5.3 Generalization across different patient populations

Generalization refers to the ability of a model trained on a particular dataset to perform well on new, unseen data. Achieving good generalization across different patient populations is crucial in medical applications to ensure the model’s reliability and effectiveness in real-world scenarios.

When dealing with ECG and PPG data, there are several challenges related to generalization across different patient populations:

#### Inter-Subject Variability

Individuals’ ECG and PPG patterns can vary significantly due to differences in age, gender, physiological characteristics, and underlying health conditions. A model trained on a specific demographic may not perform well on a different population.

#### Data Distribution Shift

If the distribution of data in the target patient population differs significantly from the training data distribution, the model may fail to generalize effectively.

#### Artifact Variability

ECG and PPG signals can be affected by artifacts like noise, motion, or electrode misplacement. These artifacts may vary across patient populations, making generalization challenging.

#### Limited Annotated Data

Obtaining labeled data for specific patient populations might be more challenging, leading to fewer samples for training and evaluation.

To improve generalization across different patient populations in self-supervised learning for ECG and PPG data, following approaches can be considered :

#### Diverse Training Data

Use a large and diverse dataset that covers a wide range of patient demographics and conditions. This can help the model learn robust features that are less sensitive to demographic variations.

#### Data Augmentation

Apply data augmentation techniques to artificially increase the diversity of the training data. This can help the model learn to be more invariant to certain types of variations.

#### Transfer Learning

Pre-train the model on a large dataset and then fine-tune it on a smaller dataset from the target population. Transfer learning can leverage knowledge learned from a larger dataset to adapt better to a specific population.

#### Domain Adaptation

Employ domain adaptation techniques to mitigate the distribution shift between the source (training) and target (testing) patient populations.

#### Cross-Validation

Perform cross-validation using multiple patient populations to assess the model’s generalization performance more comprehensively.

#### Adversarial Training

Introduce adversarial learning techniques that encourage the model to be invariant to certain variations across different patient populations.

### 5.4 Interpretability and explainability of self-supervised models

Interpretability and explainability are crucial aspects of self-supervised models for ECG (Electrocardiogram) and PPG (Photoplethysmogram) analysis. Self-supervised learning is a technique that enables models to learn from unlabeled data, making it particularly appealing for medical applications where labeled data is often scarce and costly to obtain. While self-supervised models have shown promising results in ECG and PPG tasks, their interpretability and explainability remain important challenges. Understanding the decisions and predictions made by these models is critical in medical settings, where clinicians need to trust and comprehend the reasoning behind the model’s outputs. Researchers and developers are actively exploring methods to make self-supervised models more interpretable and explainable, using techniques such as attention mechanisms, saliency maps, and feature visualization to shed light on the model’s internal workings and provide insights into how it processes and interprets ECG and PPG data. By enhancing the interpretability and explainability of self-supervised models, we can foster greater trust and adoption of these advanced techniques in clinical settings, ultimately improving patient care and outcomes.

### 5.5 Potential applications in personalized medicine and remote healthcare

Self-supervised models for electrocardiogram (ECG) and photoplethysmogram (PPG) data have the potential to revolutionize personalized medicine and remote healthcare. ECG and PPG are valuable physiological signals that provide crucial insights into a person’s cardiovascular health. By leveraging self-supervised learning, these models can learn representations directly from the raw data without the need for labeled datasets, making them adept at extracting complex patterns and correlations from large-scale, unannotated patient records. In personalized medicine, such models can aid in early detection and prediction of cardiac abnormalities, enabling healthcare providers to offer targeted interventions and personalized treatment plans based on a patient’s unique cardiac profile. Additionally, in the realm of remote healthcare, these models can empower wearable devices and mobile applications to continuously monitor individuals’ cardiovascular health in real-time, alerting them and their caregivers to any concerning deviations from their baseline. This proactive approach can lead to timely interventions and prevent adverse events, enhancing patient outcomes and overall healthcare efficiency. Furthermore, the anonymized and aggregated data from these models can be harnessed for population-wide research and insights, fostering advancements in cardiac healthcare on a global scale.

### 5.6 Emerging trends and future research directions

Emerging trends and future research directions in self-supervised models for ECG and PPG (Electrocardiogram and Photoplethysmogram) signal analysis hold tremendous potential for advancing the field of cardiovascular health monitoring and diagnosis. Self-supervised learning has shown promising results by leveraging the vast amounts of unlabeled data to pretrain deep neural networks. One key trend is the integration of multimodal data, combining ECG and PPG signals, along with other physiological measurements, to enhance model performance and provide a more comprehensive understanding of cardiovascular health. Additionally, there is a growing interest in exploring self-supervised models for anomaly detection and prediction in long-term continuous monitoring scenarios, where the models can learn to recognize deviations from individual baseline patterns, enabling timely interventions. Furthermore, the development of explainable and interpretable self-supervised methods is a crucial direction to gain insights into the decision-making process of the models, thereby increasing their clinical applicability and trustworthiness. As the availability of large-scale ECG and PPG datasets continues to grow, future research in this area should focus on designing novel self-supervised architectures, leveraging attention mechanisms, transformer-based models, and other cutting-edge techniques to further improve the accuracy, robustness, and scalability of self-supervised models for ECG and PPG signal analysis, ultimately leading to significant advancements in cardiovascular healthcare.

## 6 Conclusion

In conclusion, self-supervised models have emerged as a promising approach for the analysis of ECG and PPG signals, offering a novel and effective means of leveraging vast amounts of unlabeled data for training. These models have demonstrated their capability to autonomously learn meaningful representations from raw physiological data without the need for manual annotations, thus overcoming the limitations associated with traditional supervised learning paradigms. By exploiting the inherent structure and temporal dependencies present in ECG and PPG signals, self-supervised models have shown remarkable potential in tasks such as abnormality detection, heart rate estimation, and disease classification. Moreover, their ability to generalize across different datasets and patient populations indicates their versatility and robustness. As the field of self-supervised learning continues to evolve, the ongoing research and development in this area are likely to unlock further advancements in the medical domain, paving the way for more accurate and personalized healthcare solutions based on physiological data analysis. However, challenges remain, including data scarcity, interpretability, and model complexity, which demand further investigation to ensure the responsible and ethical deployment of self-supervised models in real-world clinical settings. Nevertheless, the strides made so far in leveraging self-supervised approaches for ECG and PPG analysis underscore their potential as a transformative force in healthcare, empowering clinicians with more sophisticated tools to enhance patient care and diagnosis.

### 6.1 Summary of the key findings and contributions

Self-supervised learning has emerged as a powerful approach for leveraging unlabeled data to train machine learning models. In the context of electrocardiogram (ECG) and photoplethysmogram (PPG) data, researchers have made significant strides in harnessing self-supervised learning techniques to extract meaningful features and improve various cardiovascular-related tasks. Here are the key findings and contributions in this domain:

#### Unsupervised Representation Learning

Self-supervised learning methods have shown promise in learning rich and informative representations from ECG and PPG signals without the need for labeled annotations. These learned representations can capture intricate patterns and relationships present in the data, aiding subsequent tasks.

#### Anomaly Detection

Self-supervised learning has been employed for anomaly detection in ECG and PPG data. By training models on a pretext task, such as predicting missing segments or reconstructing the input data, they can effectively identify abnormal heart rhythms or irregularities, which is crucial for early detection of cardiovascular diseases.

#### Transfer Learning

Self-supervised pretraining has been explored to facilitate transfer learning in the medical domain. By training a model on a large-scale dataset using self-supervised methods, researchers can fine-tune the model on smaller labeled datasets for specific tasks, such as arrhythmia classification or heart rate estimation, achieving improved performance.

#### Domain Adaptation

Self-supervised learning has also been applied to domain adaptation tasks, where models are trained on a source domain with abundant data and then adapted to perform well on a target domain with limited or different data distributions. This approach has shown promise in addressing issues related to data scarcity and domain shift in ECG and PPG analysis.

#### Data Augmentation

Self-supervised learning provides a novel way to generate augmented data for training deep learning models. By utilizing pretext tasks like temporal shuffling or sequence prediction, the models learn to extract temporal dependencies and spatial information from the ECG and PPG signals, thereby enhancing generalization and robustness.

#### Interpretability and Explainability

Self-supervised learning has also been utilized to improve model interpretability and explainability. By learning meaningful representations, researchers have been able to identify salient features in ECG and PPG data that contribute to model decisions, aiding clinicians in understanding the underlying mechanisms behind the model’s predictions.

In conclusion, self-supervised learning has shown great promise in the domain of ECG and PPG data analysis. By leveraging unlabeled data and pretext tasks, researchers have made significant contributions to representation learning, anomaly detection, transfer learning, domain adaptation, data augmentation, and model interpretability. These advancements hold the potential to enhance the accuracy and effectiveness of cardiovascular diagnosis and monitoring, ultimately benefiting patient care and treatment. However, as the field is still relatively new, continued research and exploration are necessary to fully unlock the potential of self-supervised learning in this domain.

### 6.2 Potential impact of self-supervised learning on biomedical signal analysis

Self-supervised learning has the potential to revolutionize biomedical signal analysis, particularly in the domain of electrocardiogram (ECG) and photoplethysmogram (PPG) data. By leveraging large amounts of unlabeled data, self-supervised learning algorithms can automatically learn meaningful representations from raw signals, capturing complex patterns and dependencies without the need for extensive manual annotations. This approach has the potential to enhance the accuracy and efficiency of various tasks, such as heartbeat classification, arrhythmia detection, and physiological parameter estimation. Furthermore, self-supervised learning could enable the transfer of knowledge from one domain to another, allowing models trained on ECG data to generalize effectively to PPG data, and vice versa. This not only saves data collection effort but also enhances the adaptability of the algorithms to new, unseen scenarios. Overall, the integration of self-supervised learning into biomedical signal analysis holds great promise for advancing the understanding and diagnosis of cardiovascular health, paving the way for more accessible and accurate healthcare solutions.

### 6.3 Final remarks and call for further research in this area

In conclusion, self-supervised learning leverages large amounts of unlabeled data to pretrain models and has led to improved feature representations and enhanced performance in various tasks, such as arrhythmia detection and cardiovascular risk assessment. However, further research is needed to address challenges such as data scarcity, domain adaptation, and model generalization. Additionally, exploring novel self-supervised learning techniques tailored specifically for ECG and PPG signals could unlock greater potential for enhancing healthcare diagnostics and personalized monitoring, paving the way for more accurate and efficient patient care.

## Data Availability

All data produced in the present work are contained in the manuscript

